# Multidrug-Resistant ESKAPEEc Pathogens from Bloodstream Infections in South Africa: A Cross-Sectional Study Assessing Resistance to WHO AWaRe Antibiotics

**DOI:** 10.1101/2025.04.17.25325933

**Authors:** Bakoena A. Hetsa, Jonathan Asante, Daniel G. Amoako, Akebe L. K Abia, Joshua Mbanga, Sabiha Y. Essack

## Abstract

**Background and Aims:** Multidrug-resistant (MDR) pathogens, particularly members of the ESKAPE group and *Escherichia coli* (collectively referred to as ESKAPEEc), are major contributors to bloodstream infections (BSIs) and pose significant treatment challenges. This study aimed to characterize the antimicrobial resistance (AMR) profiles of ESKAPEEc isolates from BSIs in public hospitals in the uMgungundlovu District, South Africa, and to assess their resistance to World Health Organization (WHO) Access, Watch, and Reserve (AWaRe) antibiotics.

**Methods:** Between November 2017 and December 2018, blood samples (n = 195) were collected from adult and paediatric patients with suspected BSIs. Isolates were identified using the VITEK 2® system and confirmed by polymerase chain reaction (PCR). Antimicrobial susceptibility testing was performed using the Kirby–Bauer disk diffusion method and interpreted according to EUCAST/CLSI guidelines. The multiple antibiotic resistance index (MARI) was calculated. One-way analysis of variance (ANOVA) was used to assess associations between MARI and clinical variables, including ward type and facility level

**Results:** Out of 195 presumptive isolates, 159 were confirmed as ESKAPEEc. The most frequently identified pathogens were *Klebsiella pneumoniae* (28.9%) and *Staphylococcus aureus* (28.3%). High resistance rates were observed across WHO Access and Watch antibiotics, including ampicillin (76% in *E. coli*), gentamicin (67.4% in *K. pneumoniae*), and ciprofloxacin (≥60% in most species). Carbapenem resistance in *Acinetobacter baumannii* reached 90%. Overall, 94.9% of isolates were MDR, and 93.1% had MARI ≥0.2. Significant differences in MARI values were observed across ward groups and facility levels, with the highest values recorded in intensive care units (mean = 0.67, 95% CI: 0.62–0.72) and tertiary hospitals (mean = 0.64, 95% CI: 0.60–0.68), compared to regional hospitals (mean = 0.52, 95% CI: 0.47–0.57)

**Conclusion:** The findings reveal a high burden of MDR ESKAPEEc in BSIs and widespread resistance to WHO Watch antibiotics. Targeted antimicrobial stewardship and the implementation of microbiology-guided therapy are urgently needed to optimize patient outcomes and curb the spread of resistance.

## 1.0 Introduction

*Enterococcus faecium*, *Staphylococcus aureus*, *Klebsiella pneumoniae*, *Acinetobacter baumannii*, *Pseudomonas aeruginosa* and *Enterobacter* spp. are called ESKAPE pathogens that escape the action of antibiotics due to their increasing multidrug resistance (De Socio et al., 2019; Santajit and Indrawattana, 2016). Together with third-generation cephalosporin and carbapenem-resistant *Escherichia coli*, these bacteria are identified by the World Health Organization as bacterial priority pathogens for the research and development of new antibiotics (WHO, 2024). The ESKAPE and *E. coli* pathogens (ESKAPEEc) are increasingly implicated in most nosocomial infections, including bloodstream infections (BSIs) and sepsis (De Angelis *et al*., 2018). These infections are commonly reported among neonates, the elderly, and immunocompromised patients (Martinez and Wolk, 2016).

BSIs are invasive infections characterized by high mortality rates and healthcare costs (Rudd *et al*., 2020). Several studies on BSIs have reported different causative organisms with varying occurrences of Gram-negative (Musicha *et al*., 2017) and Gram-positive (Crichton *et al*., 2018; Mpinda-Joseph et al., 2019) pathogens. A 20-year surveillance study by Tian *et al*. (2019) in China found that *E. coli*, *S. aureus,* and *K. pneumoniae* were the most predominantly identified pathogens causing BSIs. Their results revealed that *K*. *pneumoniae* and *E. coli* displayed resistance to antibiotics, including third-generation cephalosporins and fluoroquinolones (Tian, Zhang and Sun, 2019).

In South Africa, carbapenem-resistant *Enterobacterales* (CRE) have been reported in a retrospective survey done in patients with bacteraemia from 2015 to 2018 through the Group for Enteric, Respiratory and Meningeal Disease Surveillance in South Africa (GERMS-SA) surveillance platform. Perovic *et al*. (2020a) revealed that more than 75% of CRE isolates were *K. pneumoniae* and carried *bla_OXA-48_* and *bla_NDM_* genes conferring resistance to carbapenems. Moreover, a more recent study that compared the BSI incidence, pathogens, and antimicrobial resistance profiles in two large neonatal units in Botswana and South Africa showed that *Klebsiella pneumoniae* and *S. aureus* were dominant pathogens and displayed high levels of resistance to antibiotics (Gezmu *et al*., 2021). Another South African study on ESKAPEEc isolated from patients with bacteraemia in private and public sectors/hospitals identified *K. pneumoniae* as the leading pathogen. Isolates displayed varying non-susceptibility to ciprofloxacin, cephalosporins, carbapenems, ciprofloxacin, amikacin, gentamicin, and tigecycline (Ismail and Perovic, 2018).

The surveillance and characterization of antibiotic-resistant pathogens, including ESKAPEEc, is essential in managing the rise of AMR. We assessed the antibiotic resistance profiles of ESKAPEEc implicated in BSIs among adults and children in two public hospitals in the uMgungundlovu district in KwaZulu-Natal, South Africa.

## 2.0 Materials and Methods

### 2.1. Ethical approval

Ethical approval was obtained from the Biomedical Research Ethics Committee of the University of KwaZulu-Natal (Reference Number: BCA444/16). All bacterial isolates were obtained as part of routine clinical diagnostics, and no patient-identifiable information was collected. As such, the requirement for individual informed consent was waived by the ethics committee.

### 2.2. Study setting and sample collection

Blood cultures were collected from November 2017 to December 2018 at one regional and one tertiary public sector hospitals in the uMgungundlovu District. The regional hospital comprised 824 beds, while the tertiary hospital is a 530-bed facility. A total of 195 presumptive ESKAPEEc isolates implicated in BSIs constituted the sample. Patients’ demographic data (age, sex, ward type, and specimen source) were collected. Patients’ names were excluded to maintain anonymity.

### 2.3. Bacterial Isolation and Identification

Preliminary identification of isolates was done using the VITEK 2^®^ system (BioMérieux, Marcy-L’Etoile, France). Pure isolates were stored in tryptic soy broth (Basingstoke, Hampshire, England) broth supplemented with 20% glycerol at −80 °C until further analysis.

### 2.4 DNA extraction and molecular confirmation of ESKAPE and E. coli isolates

DNA was extracted using the boiling method as previously described (Chen et al., 2011). The purity and concentration of extracted DNA were determined by gel electrophoresis. Molecular confirmation of ESKAPEEc isolates was done using species-specific oligonucleotide primer sequences and PCR conditions listed in **Table S2**. PCR was performed in a 10 µL reaction volume with 3 µL DNA template, 5 µL Luna® Universal qPCR master mix (Biolabs, New England, South Africa), 0.5 mL (1.25 µM) of each primer, and 1 mL of nuclease-free water. *E. coli* ATCC 25922, *S. aureus* ATCC 29213, *K. pneumoniae* ATCC 35657, *E. aerogenes* ATCC 13048, *Enterococcus* spp. ATCC 29212, *P. aeruginosa* ATCC 35032, and *A. baumannii* ATCC 19606 were used as positive controls, while nuclease-free water was used as negative control.

### 2.5 Molecular Identification of Methicillin-Resistant S. aureus (MRSA)

Multiplex PCR was conducted targeting *S. aureus* specific thermonuclease (*nuc)* gene, and the methicillin-resistance-encoding *mecA* gene using primers and PCR conditions shown in Table S2. All reactions were performed in a total volume of 10 uL, which consisted of 5 µL 2× Luna^®^ Universal qPCR Master Mix (New England Biolabs, South Africa) 0.5 µL (1.25 mM) of the forward and reverse primers, 1 µL of nuclease-free water, and 3 µL of template DNA. The PCR conditions were as previously described (Amoako *et al*., 2019), with slight modifications of the hot start activation (94 °C for 5 min), followed by 35 cycles of denaturation (94 °C for 30s), and annealing/extension at (62 °C for 30s). A melt curve was prepared by ramping up the melting temperature from 60 °C to 95 °C at a rate of 0.15 °C/s on a continuous mode following a pre-melt step at 95 °C for 15 s on the 1st step. The methicillin-resistant *S. aureus* ATCC 43300, was used as a positive control, and nuclease-free water was used as the negative control. All reactions were carried out in an Applied Biosystems QuantStudio 5 Real-time PCR system (Thermo Fisher Scientific, Waltman, MA).

### 2.6. Antimicrobial Susceptibility Testing (AST)

The antibiotic susceptibility profiles were determined by the Kirby–Bauer disk diffusion method, and interpreted according to the European Committee on Antimicrobial Susceptibility Testing (EUCAST) breakpoints (EUCAST, 2017). Breakpoints for ampicillin, azithromycin, tetracycline, and nalidixic acid were from the Clinical and Laboratory Standards Institute (CLSI) breakpoints (CLSI., 2017). Antibiotic resistance profiles were classified using the WHO AWaRe arrangement of Access (first line), Watch (restricted), Reserve (last resort) antibiotics (WHO AWaRe, 2023). For *Enterobacterales* the following 20 antibiotics were used: amikacin (AMK,30 µg), ampicillin (AMP, 10 µg), azithromycin (AZM, 15 µg), amoxicillin-clavulanic acid (AMC, 30 µg), cefepime (FEP, 10 µg), cefotaxime (CTX, 30), cefoxitin (FOX, 30 µg), ceftazidime (CAZ, 30 µg), ceftriaxone (CRO, 30 µg), cephalexin (LEX, 30 µg), ciprofloxacin (CIP, 5 µg), chloramphenicol (CHL, 30 µg), gentamicin (GEN, 10 µg), imipenem (IPM, 10 µg), meropenem (MEM, 10 µg), nalidixic acid (NAL, 30 µg), piperacillin-tazobactam (TZP, 110 µg), tetracycline (TET, 30 µg), tigecycline (TGC, 15 µg), and trimethoprim-sulfamethoxazole (SXT, 25 µg) (Oxoid, Basingstoke, UK), were used. All antibiotic discs used in this study were purchased from Oxoid (Oxoid Ltd., Basingstoke, UK) (Table S 1). *E. coli* ATCC 25922 was used for quality control.

For *E. faecium*, 16 commercial antibiotic disks were tested, which included ampicillin (AMP, 10 µg), chloramphenicol (CHL, 30 µg), ciprofloxacin (CIP, 5 µg), erythromycin (ERY, 15 µg), imipenem (IPM, 10 µg), gentamycin (GEN, 120 µg), levofloxacin (LVX, 5 µg), linezolid (LZD, 30 µg), tetracycline (TET, 30 µg), nitrofurantoin (NIT, 300 µg), quinupristin-dalfopristin (Q-D, 15 µg), streptomycin (STR, 300 µg), teicoplanin (TEC, 30 µg), tigecycline (TGC, 15 µg), trimethoprim-sulfamethoxazole (SXT, 25 µg), and vancomycin (VAN, 30 µg), disks (Oxoid, Basingstoke, UK), (interpreted using EUCAST breakpoint (EUCAST, 2019). Breakpoints for chloramphenicol (CHL 30 µg), tetracycline (TET, 30 µg), and erythromycin (ERY, 15 µg) were interpreted according to CLSI (CLSI., 2017). *E. faecalis* ATCC 29212 was used for quality control.

For *S. aureus* the following 20 antibiotics were used: penicillin G (PEN 10 µg), ampicillin (AMP 10 µg), amikacin (AMK, 30 µg), cefoxitin (FOX, 30 µg), chloramphenicol (CHL, 30 µg), clindamycin (CLI, 2 µg), ciprofloxacin (CIP, 5 µg), erythromycin (ERY, 15 µg), gentamicin (GEN, 10 µg), levofloxacin (LVX, 5 µg), linezolid (LZD 30 µg), moxifloxacin (MXF, 5 µg), quinupristin-dalfopristin (Q-D, 15 µg), rifampicin (RIF, 5 µg), tetracycline (TET, 30 µg), tigecycline (TGC, 15 µg), trimethoprim-sulfamethoxazole (SXT, 25 µg), (interpreted using EUCAST breakpoints), and doxycycline (DOX, 30 µg), nitrofurantoin (NIT, 300µg), teicoplanin (TEC, 30 µg) interpreted using CLSI breakpoints. All antibiotics were purchased from Oxoid (Oxoid, Basingstoke, UK). The minimum inhibitory concentrations (MICs) for vancomycin (VAN) were determined through the broth microdilution method using the CLSI guidelines, due to the absence of breakpoints for the disc diffusion method (CLSI., 2017). A methicillin-sensitive strain, *S. aureus* ATCC 29213, was used as a positive control.

### 2.7 Multidrug-Resistance and Multiple Antimicrobial Resistance Index (MARI) Analysis

Multidrug resistance was defined as resistance to at least one agent in three distinct antibiotic classes. Multiple antibiotic resistance phenotypes were displayed by all isolates in the study regardless of antibiotic classes. The MAR index was evaluated using the formula MAR=x/y, where x is the number of antibiotics the isolate was resistant to, and y is the total number of antibiotics tested against the isolate (Krumperman, 1983). The MAR index was used as an indicator of health risk assessment to ascertain whether isolates originate from excessive or low antibiotic use environments.

### 2.8. Statistical Analysis

All statistical analyses were performed using IBM SPSS Statistics for Windows, version 27.0 (IBM Corp., Armonk, NY, USA) and Python 3.11 with the statsmodels and seaborn libraries for data visualization. Descriptive statistics were used to summarize demographic characteristics and antimicrobial resistance data. Continuous variables were presented as means with standard deviations (SD) or medians with interquartile ranges (IQR), where appropriate. Categorical variables were reported as frequencies and percentages.

The multiple antibiotic resistance index (MARI) was calculated for each isolate by dividing the number of antibiotics to which the isolate was resistant by the total number of antibiotics tested. One-way analysis of variance (ANOVA) was used to compare MARI values across two key categorical variables: facility level (regional vs. tertiary hospitals) and clinical setting, classified into Ward Group categories (Intensive Care Units, General Inpatient Wards, Surgical Wards, Paediatric Units, Emergency/Trauma Units, Specialist Wards, and Outpatient/Clinic-Based Services).

The assumptions of normality and homogeneity of variances were assessed using the Shapiro– Wilk and Levene’s tests, respectively. Where these assumptions were met, ANOVA was conducted to identify overall group differences. Tukey’s Honestly Significant Difference (HSD) test was used post hoc to identify specific group pairwise differences. Effect sizes were reported using partial eta squared (η²) to quantify the proportion of variance in MARI explained by each factor. Confidence intervals (95% CI) for group means were calculated to assess the precision of the estimates. All statistical tests were two-sided, and a significance level (α) of 0.05 was used to determine statistical significance. No imputation was performed for missing data. All analyses were pre-specified in the study protocol, and no exploratory subgroup analyses were conducted. Statistical terminology, abbreviations, and test types were clearly defined in the results section to ensure interpretability and consistency.

## 3. Results

### 3.1. Isolation, Molecular Identification and Distribution of Isolates

The VITEK 2 system was utilized for bacterial identification. Of the 185 presumptive isolates, 159 (85.9%) were confirmed as members of the ESKAPEEc group using the VITEK 2 system. Species-level identification was further validated using qPCR with species-specific oligonucleotide primers listed in Supplementary Table S1. No discrepancies were observed between phenotypic and molecular identification across all isolates. Among the ESKAPEEc pathogens, *Klebsiella pneumoniae* was the most frequently isolated species (n = 46, 28.9%), followed closely by *Staphylococcus aureus* (n = 45, 28.3%), and *Escherichia coli* (n = 25, 15.7%) (Figure 1A). Twenty-six (26) isolates were identified as non-ESKAPEEc organisms, including *Enterococcus faecalis* (n = 13, 50.0%), *Serratia marcescens* (n = 11, 42.3%), and *Proteus mirabilis* (n = 2, 7.6%).

**Figure 1.**
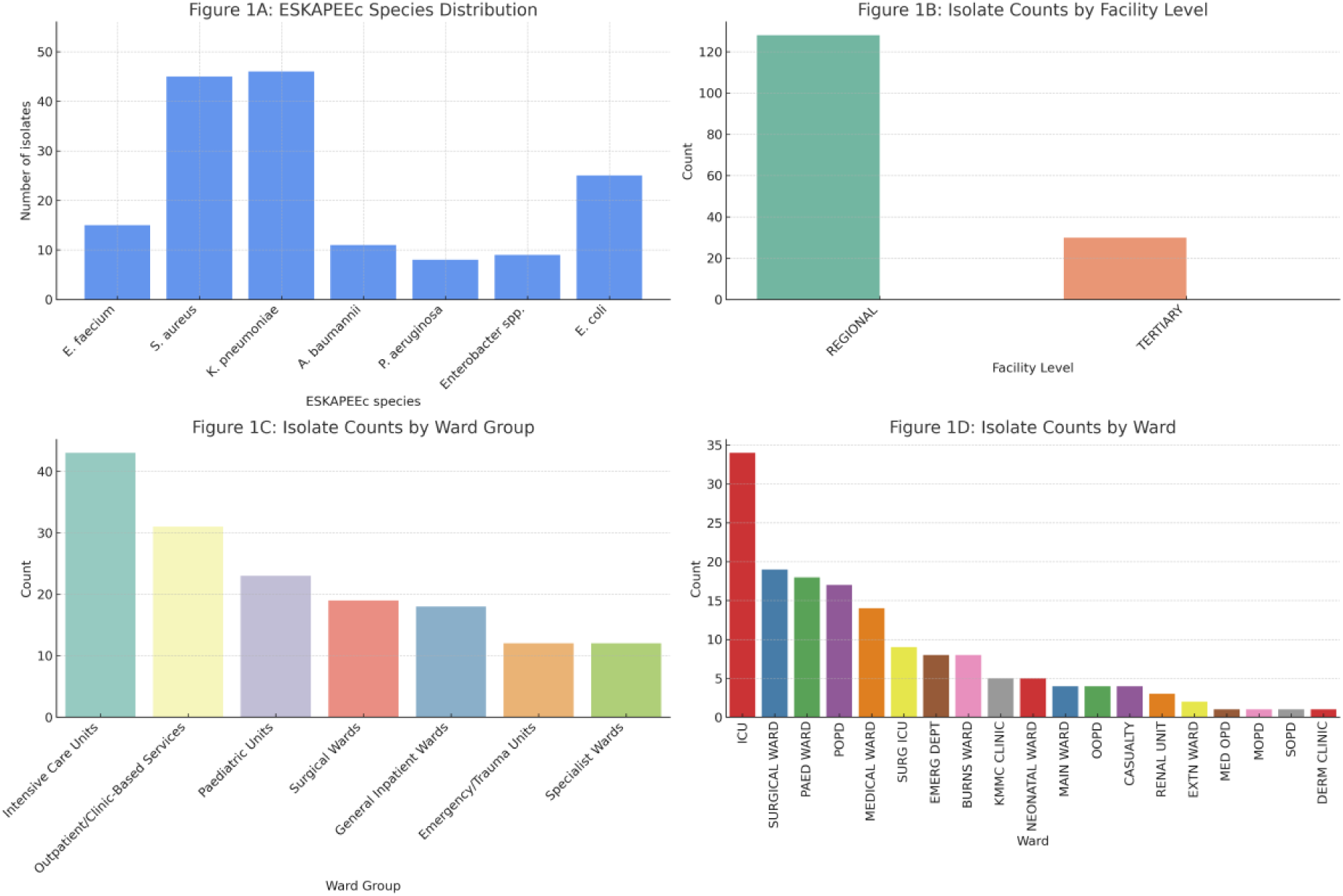
Distribution of ESKAPEEc Isolates by Species, Facility Level, Ward Group, and Ward Type. (A) Frequency distribution of ESKAPEEc species isolated from bloodstream infections (n =159). *Klebsiella pneumoniae*and *Staphylococcus aureus* were the most common species. (B) Isolate counts stratified by healthcare facility level, showing a predominance of isolates from regional hospitals compared to tertiary hospitals. (C) Isolate distribution across broader clinical categories (WardGroup), with Intensive Care Units (ICUs) and Outpatient/Clinic-Based Services accounting for the highest numbers. (D) Detailed breakdown of isolate counts by specific hospital wards, with ICU, surgical, and pediatric wards contributing the highest proportions.

Regarding facility level distribution, the majority of isolates were obtained from regional hospitals (n = 128, 80.5%), while tertiary hospitals contributed 31 isolates (19.5%) (Figure 1B). Stratification by broader clinical settings (Ward Group) revealed that the highest number of isolates originated from Intensive Care Units (n = 43), followed by Outpatient/Clinic-Based Services (n = 31), and Paediatric Units (n = 24) (Figure 1C). Across clinical departments, the Intensive Care Unit (ICU) accounted for the largest share of isolates (n = 33, 20.7%), followed by surgical and paediatric wards (Figure 1D). Within the ICU, *K. pneumoniae* was the predominant pathogen (15/33, 45.5%), while *S. aureus* was most frequently recovered from the paediatric outpatient department (n = 12). The remaining species were variably distributed across other ward types levels (Figures 1D; Supplementary Table S2).

Seventy-nine of the 159 patients (49.7%) were males, and 71 (44.7%) were females, while the remaining 9 (5.6%) were unidentified. The participants’ age ranged from 0 months to 77 years. The mean age was 19.35 years. Also, 3 (6.7%), of the 45 *S. aureus* isolates were confirmed as MRSA by PCR detection of *mec*A gene.

### 3.2. Prevalence of antibiotic-resistance profiles of ESKAPEEc isolates

The antibiotic susceptibility profiles of confirmed ESKAPEEc isolates are summarized in Table 1. *Klebsiella pneumoniae* exhibited the highest resistance to Access group antibiotics, notably cephalexin (78.3%), amoxicillin-clavulanic acid (69.6%), and trimethoprim-sulfamethoxazole (67.4%), with the lowest resistance to amikacin (39.1%). Resistance to Watch group antibiotics ranged from 30.4% to 69.0%, with highest resistance against cefotaxime, ceftazidime, and ciprofloxacin (each 67.4%–69.0%), and the lowest against imipenem (30.4%) and meropenem (32.6%). All *K. pneumoniae* isolates were susceptible to tigecycline (91.3%) and were not tested for ampicillin due to intrinsic resistance (Holt *et al*., 2015). *Escherichia coli* showed high resistance to ampicillin (76.0%) and amoxicillin-clavulanic acid (68.0%) in the Access group, and 68.0% resistance to Watch antibiotics such as cefepime and ceftazidime. Lowest resistance was recorded for imipenem and meropenem (both 12.0%).

**Table 1:**
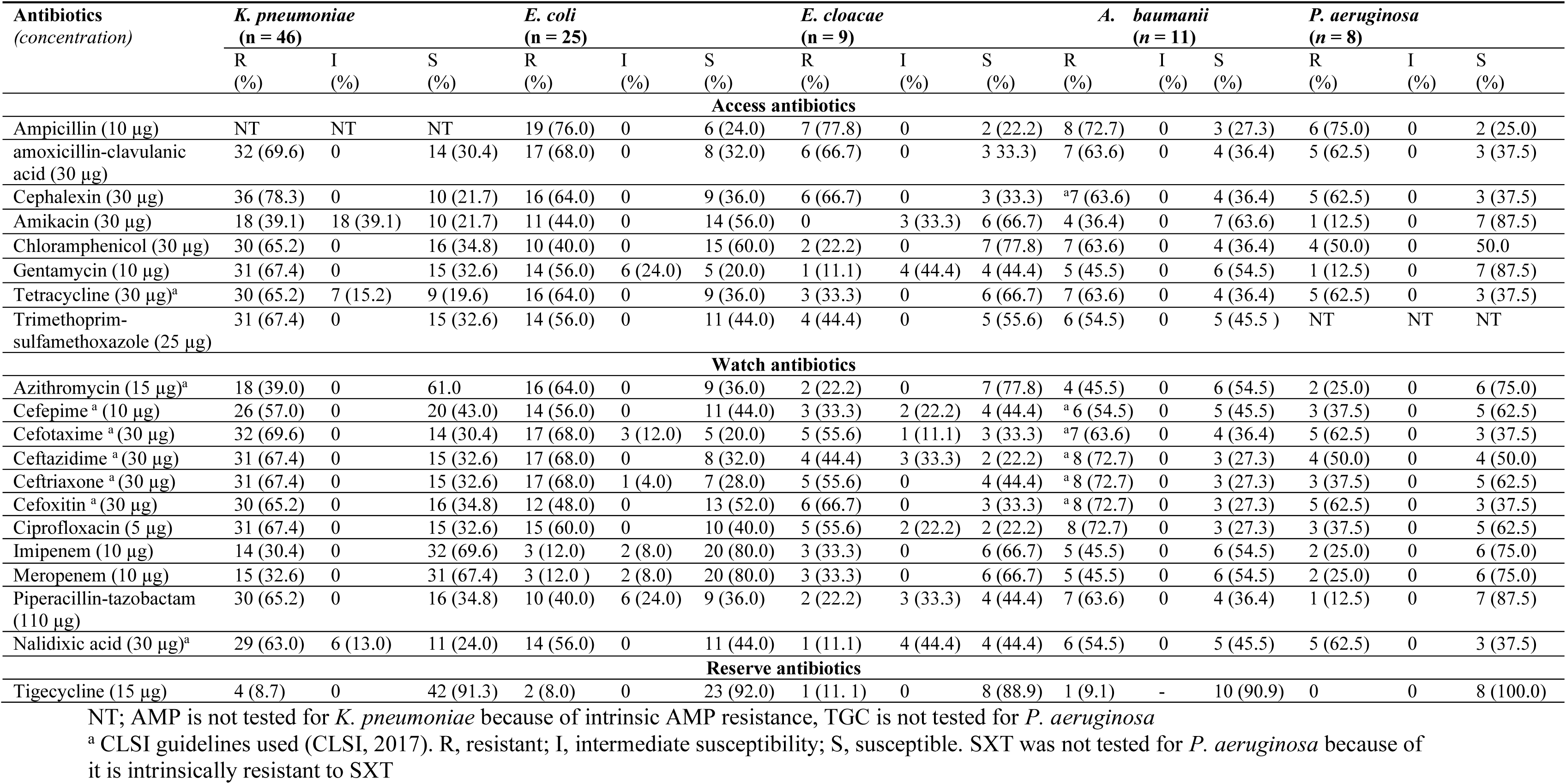
Antimicrobial susceptibility of confirmed *Enterobacterales*, and non-fermenting Gram-negative isolates from blood samples.

*Enterobacter cloacae* displayed high resistance to ampicillin (77.8%) and cefoxitin (66.7%), while resistance was lowest for gentamicin and nalidixic acid (11.1%). Isolates were highly susceptible to tigecycline (88.9%). *Acinetobacter baumannii* showed extensive resistance to several Access antibiotics including ampicillin (72.7%) and chloramphenicol (63.6%), and to Watch antibiotics such as ceftazidime, ceftriaxone, and ciprofloxacin (each 72.7%). Lowest resistance was observed for amikacin (36.4%), azithromycin (45.5%), imipenem (45.5%), and meropenem (45.5%). Tigecycline remained highly effective (90.9% susceptibility). *Pseudomonas aeruginosa* isolates were highly resistant to Access antibiotics such as ampicillin (75.0%) and amoxicillin-clavulanic acid (62.5%), but showed lower resistance to chloramphenicol and gentamicin (both 12.5%). In the Watch group, resistance was highest to cefoxitin and cefotaxime (62.5%) and lowest for piperacillin-tazobactam (12.5%). Tigecycline was not tested due to known ineffectiveness against *P. aeruginosa* (Table 1).

The antibiotic susceptibility profiles of all confirmed *S. aureus* and *E. faecium* isolates are presented in Table 2. *S. aureus* exhibited high resistance to Access group antibiotics, particularly penicillin G (93.3%) and tetracycline (60.0%), with the lowest resistance observed for chloramphenicol (17.8%). Among Watch group antibiotics, notable resistance was seen against rifampicin (62.2%), moxifloxacin (60.0%), and ciprofloxacin (57.8%). In contrast, isolates were highly susceptible to Reserve antibiotics including linezolid (95.6%), tigecycline (93.3%), and quinupristin-dalfopristin (93.3%). *E. faecium* isolates also showed substantial resistance to Access antibiotics, including ampicillin (80.0%), tetracycline (66.7%), and trimethoprim-sulfamethoxazole (66.7%). Resistance in the Watch category was recorded against erythromycin and streptomycin (both 66.7%), with the lowest resistance observed for imipenem (33.3%). Reserve agents such as vancomycin (93.3%), linezolid (96.0%), tigecycline (93.3%), and quinupristin-dalfopristin (93.3%) remained highly effective. The activity of trimethoprim and SXT against enterococci is uncertain; hence, the wild-type population is categorized as intermediate (Molechan *et al*., 2019). Consequently, there were no susceptible isolates (Table 2).

**Table 2:** Antimicrobial susceptibility of confirmed *S. aureus* and *E. faecium* isolates.

| Antibiotics (concentration) | <u>*S. aureus* (n=45)</u> |  |  | <u>*E. faecium* (n=15)</u> |  |  |
| --- | --- | --- | --- | --- | --- | --- |
|  | R (%) | I (%) | S (%) | R (%) | I (%) | S (%) |
| **Access antibiotics** |  |  |  |  |  |  |
| Penicillin G (10 µg) | 42 (93.3) | 0 | 3 (6.7) | NT | NT | NT |
| Ampicillin (10 µg) | 19 (42.2) | 0 | 26 (57.8) | 12 (80.0) | 0 | 3 (20.0) |
| Amikacin <sup>d</sup> (30µg) | 19 (42.2) | 9 (20.0) | 17 (37.8) | NT | NT | NT |
| Chloramphenicol (30 µg) | 8 (17.8) | — | 37 (82.2) | 4 (26.7) | 3 (20.0) | 8 (53.3) |
| Clindamycin <sup>d</sup> (2 µg) | 11 (24.4) | 2 (4.4) | 32 (71.2) | NT | NT | NT |
| Doxycycline <sup>d</sup> (30µg) | 13 (29.0) | 5 (11.1) | 27 (59.9) | NT | NT | NT |
| Gentamycin (10µg) | 20 (44.4) | — | 25 (55.6) | <sup>c</sup>53.3 | — | — |
| Tetracycline <sup>b</sup> (30 µg) | 27 (60.0) | — | 18 (40.0) | <sup>b</sup> 10 (66.7) | 3 (20.0) | 2 (13.3) |
| Trimethoprim-sulfamethoxazole (25µg) | 23 (51.1) | 2 (4.4) | 20 (44.5) | 10 (66.7) | 5 (33.3) | 0 |
| Nitrofurantoin <sup>a</sup> (300µg) | 12 (26.7) | 5 (11.1) | 28 (62.2) | 5 (33.3) | 3 (20.0) | 7 (46.7) |
| **Average** | **43.1%** |  |  | **54.5** |  |  |
| **Watch antibiotics** |  |  |  |  |  |  |
| Cefoxitin <sup>d</sup> (30 µg) | 17 (37.8) | — | 28 (62.2) | NT | NT | NT |
| Ciprofloxacin (5 µg) | 26 (57.8) | — | 19 (42.2) | 8 (53.3) | 0 | 7 (46.7) |
| Erythromycin <sup>a</sup> (15 µg) | 19 (42.2) | — | 26 (57.8) | 10 (66.7) | 5 (33.3) | 0 |
| Imipenem <sup>b</sup> (10 µg) | NT | NT | NT | 5 (33.3) | 2 (13.3) | 8 (53.3) |
| Levofloxacin (5µg) | 20 (44.4) | — | 25 (55.6) | 8 (53.3) | 0 | 7 (46.7) |
| Moxifloxacin (5 µg) | 27 (60.0) | — | 18 (40.0) | NT | NT | NT |
| Rifampicin <sup>d</sup> (5 µg) | 28 (62.2) | 11 (24.4) | 6 (13.3) | NT | NT | NT |
| Streptomycin <sup>c</sup> (300 µg) | NT | NT | NT | 10 (66.7) | — | — |
| Teicoplanin <sup>b</sup> (30 µg) | <sup>a</sup>18 (40.0) | 3 (6.7) | 24 (53.3) | <sup>b</sup> 6 (40.0) | — | 9 (60.0) |
| Vancomycin (30 µg)<sup>b</sup> | 0 | 0 | 45 (100.0) | 1 (6.7) | 0 | 14 (93.3) |
| **Average** | **43.1%** |  |  | **42.9%** |  |  |
| **Reserve antibiotics** |  |  |  |  |  |  |
| Linezolid (30 µg) | 2 (4.4) | — | 43 (95.6) | 1 (6.7) | — | 14 (93.3) |
| Tigecycline (15 µg) | 3 (6.7) | — | 42 (93.3) | 1 (6.7) | — | 14 (93.3) |
| Quinupristin-dalfopristin (15 µg)<sup>b</sup> | 3 (6.7) | 0 | 42 (93.3) | 2 (13.3) | — | 13 (86.7) |
| **Average** | **5.9%** |  |  | **4.9%** |  |  |

NT=Not tested PEN G, FOX, CLI, RIF, DOX, AMK
<sup>a</sup> CLSI guidelines used (CLSI, 2017).
<sup>b</sup> Based on *Enterococcus faecium* only; SXT conc (25µg).
<sup>c</sup> Only HLR considered. HLR, high-level resistance GEN (120 µg);
I, intermediate susceptibility; R, resistant; S, susceptible.
—, no breakpoints available.

Overall, high levels of resistance were observed to Access and Watch group antibiotics across the pathogens (Figure 2). *Klebsiella pneumoniae* showed the highest resistance to both Access (64.6%) and Watch (56.7%) antibiotics. *Acinetobacter baumannii* followed closely with 58.0% and 60.3% resistance to Access and Watch groups, respectively. *E. faecium*, *E. coli*, and *S. aureus* also demonstrated notable resistance rates, exceeding 40% for at least one category. In contrast, resistance to Reserve antibiotics remained relatively low across all species, with the highest being *Enterobacter cloacae* (11.1%) and *A. baumannii* (9.1%). *Pseudomonas aeruginosa* displayed no resistance to Reserve agents, though it maintained moderate resistance to Access and Watch categories (Figure 2). These findings emphasize the widespread resistance to first-line and second-line treatments

**Figure 2.**
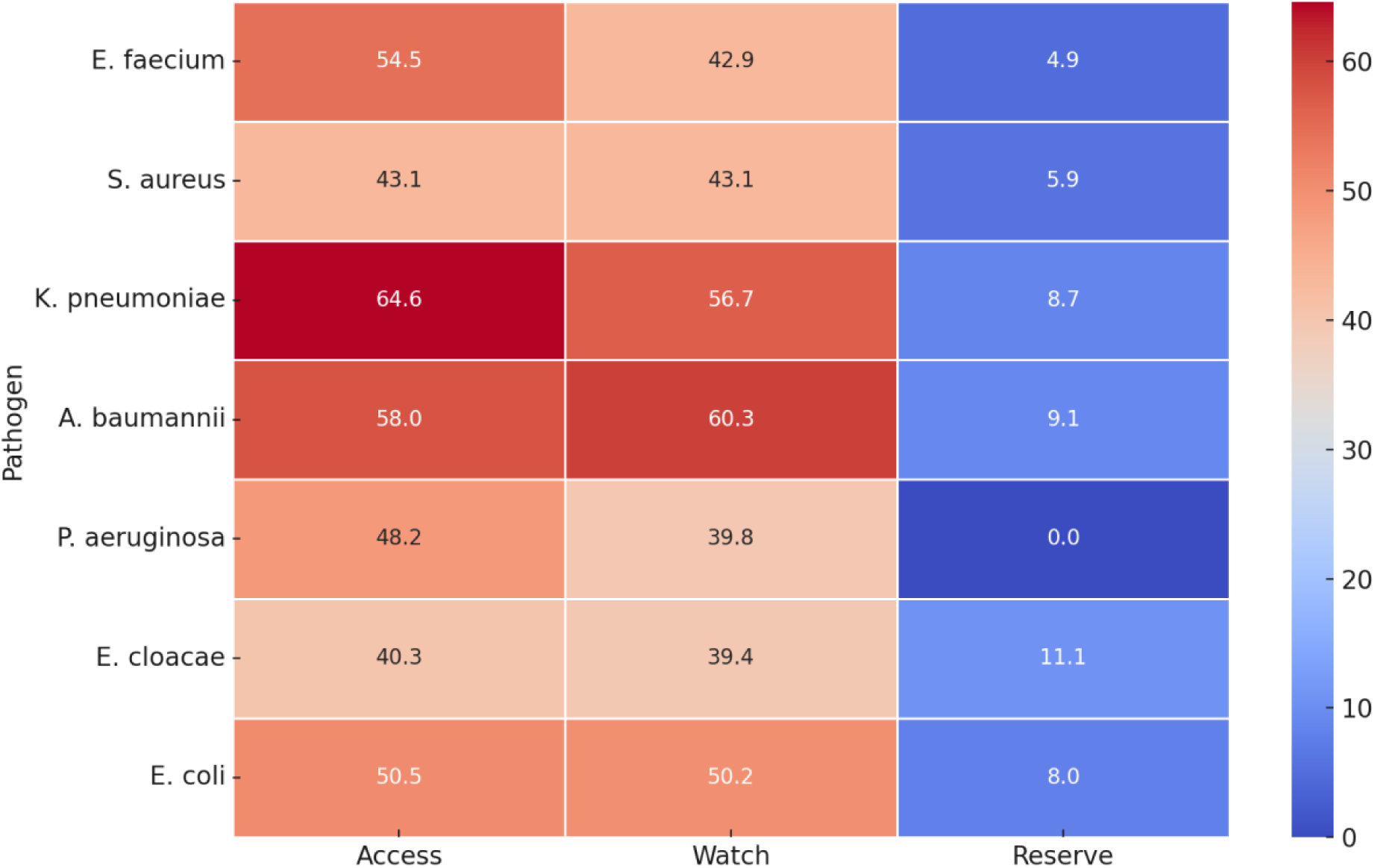
Heatmap showing the percentage of resistance to WHO AWaRe (Access, Watch, Reserve) antibiotics among ESKAPEE pathogens isolated from bloodstream infections. Resistance levels are color-coded, with higher percentages indicated by warmer tones. *Klebsiella pneumoniae* and *Acinetobacter baumannii* exhibited the highest resistance to both Access and Watch category antibiotics, while *Enterobacter cloacae* had the highest resistance to Reserve antibiotics.

#### 3.3.1 Multidrug resistance and Multiple Antibiotic Resistance (MAR) Index (MARI)

The overall MARI across all isolates ranged between 0.05 to 1.0 (mean=0.56) (Table S3). In total 148 (93.1%) had MARI **>**0.2 isolates indicating that isolates were from environments with high antibiotic use. Multidrug resistance was recorded in 151 (94.9%) isolates (Tables S4-S6). The isolates displayed varying resistance patterns that were grouped into 131 different antibiograms (Table S7).

#### 3.3.2 Statistical analysis

The multiple antibiotic resistance index (MARI) was used as a proxy for health risk assessment, providing insight into whether isolates originated from environments with high or low antibiotic pressure. To examine differences in MARI values across clinical settings, a one-way analysis of variance (ANOVA) was conducted for two grouping variables: ward group and facility level. The assumptions of normality and homogeneity of variances were evaluated prior to analysis using the Shapiro–Wilk and Levene’s tests, respectively. Both assumptions were satisfied, with Levene’s test indicating equal variances across ward groups (F = 1.001, P = .427) and facility levels (F = 3.303, P = .071) (Supplementary Table S8A).

A statistically significant difference in MARI values was observed among ward group categories (F(6, 129) = 2.895, P = .011), with a moderate effect size (η² = 0.103), suggesting that 10.3% of the variation in resistance levels was attributable to ward type (Supplementary Table S8B). The highest mean MARI values were recorded in Specialist Wards, Surgical Wards, and Intensive Care Units (Figure 3A). Post hoc analysis using Tukey’s HSD revealed a significant pairwise difference between Intensive Care Units and Outpatient/Clinic-Based Services (P = 0.031), while other pairwise comparisons were not statistically significant (Supplementary Table S8C). In addition, a significant difference was observed between regional and tertiary hospitals (F(1, 156) = 17.520, P < .001), with an effect size of η² = 0.101 (Supplementary Table S8B). Tertiary hospitals had higher average MARI scores than regional facilities, likely reflecting greater antimicrobial selection pressure and patient acuity (Figure 3B).

**Figure 3.**
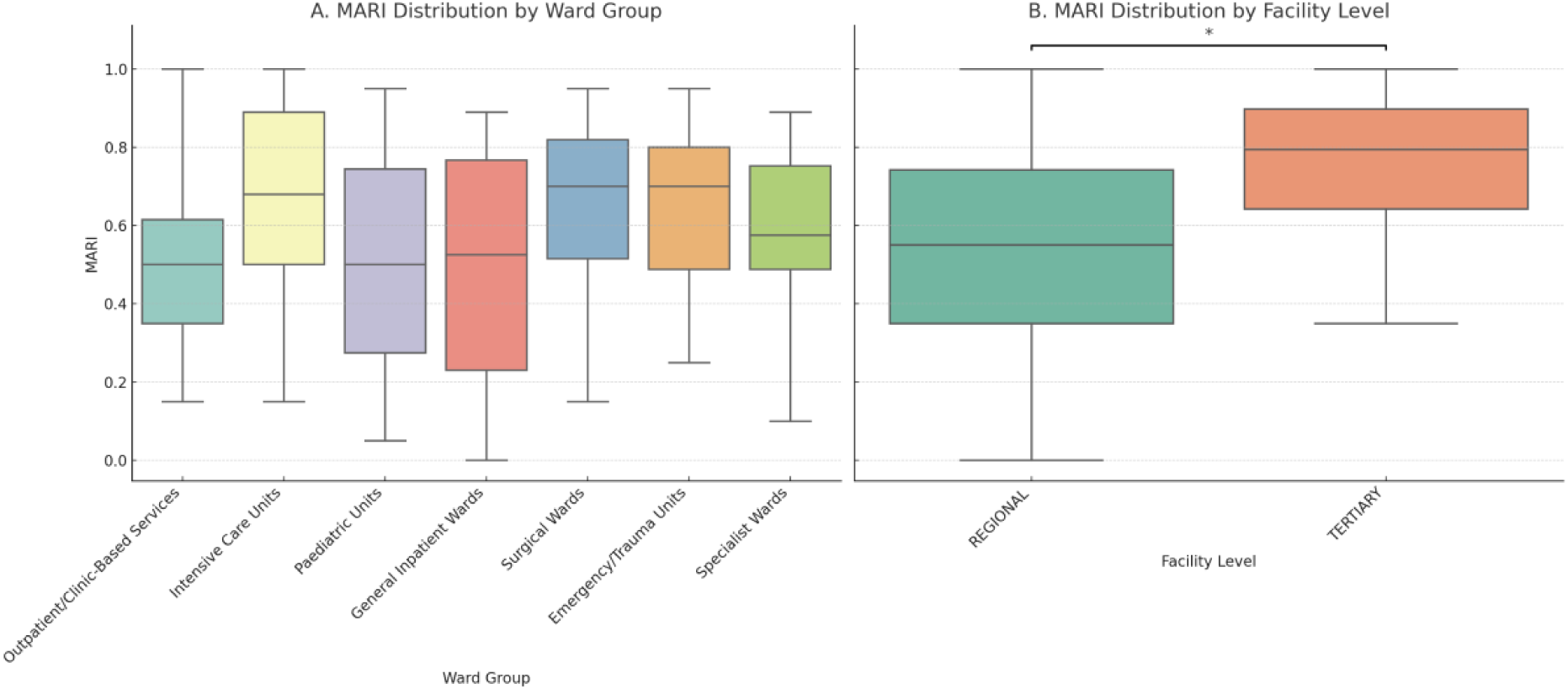
Distribution of Multiple Antibiotic Resistance Index (MARI) Values by Clinical Setting. (A) Boxplot showing the distribution of MARI values across seven clinical ward groupings. The highest median MARI scores were observed in Intensive Care Units, Surgical Wards, and Specialist Wards, while Outpatient/Clinic-Based Services had the lowest. (B) Boxplot comparing MARI values between facility levels. Tertiary hospitals showed significantly higher MARI values compared to regional hospitals (*P* < .001). The asterisk denotes a statistically significant difference.

Ninety-five percent confidence intervals (95% CI) were calculated around the group means to assess the precision of the estimates. Full descriptive statistics for both ward groups and facility levels are provided in Supplementary Tables 910A and 910B, respectively. Visual comparisons of MARI distribution and mean values are presented in Figure 3 (boxplots with significance markers) and Figure 4 (point plots with 95% confidence intervals and annotated sample sizes).

**Figure 4.**
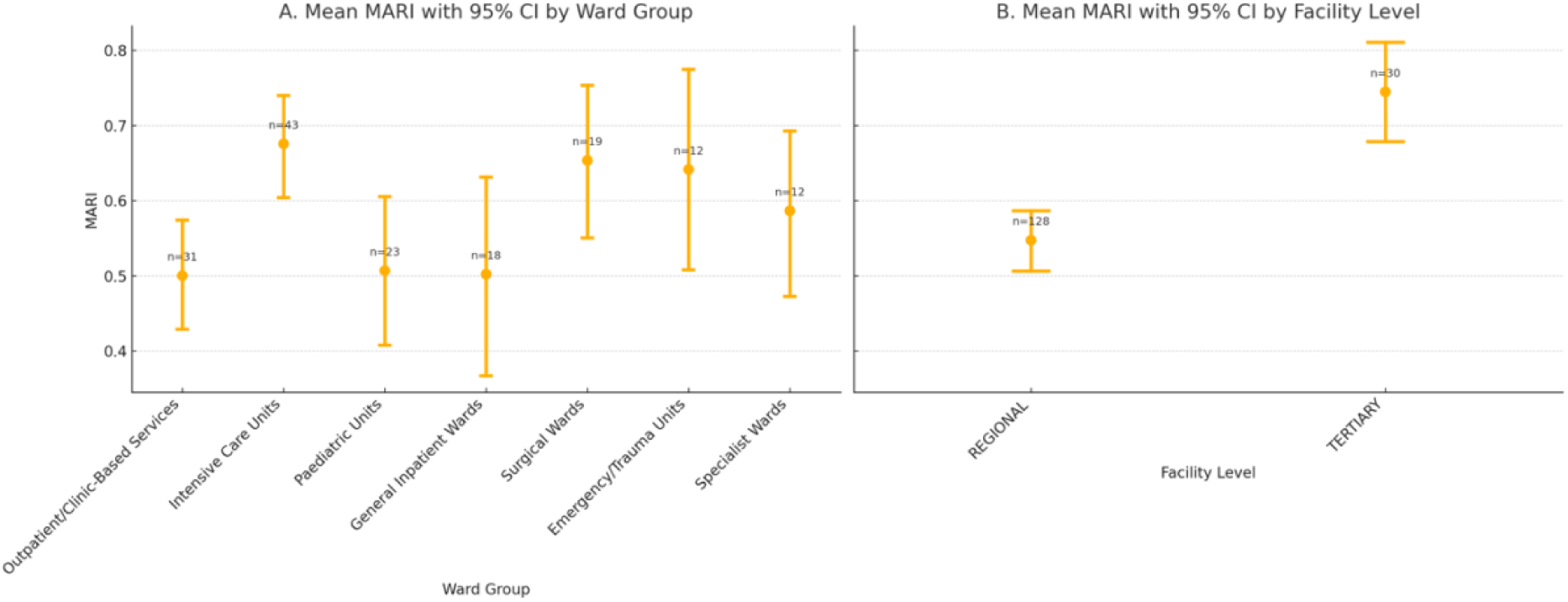
Mean Multiple Antibiotic Resistance Index (MARI) with 95% Confidence Intervals by Clinical Setting. (A) Mean MARI values with 95% confidence intervals across seven ward group categories. Intensive Care Units, Surgical Wards, and Specialist Wards exhibited the highest mean MARI values. Sample sizes for each group are annotated above the error bars. (B) Mean MARI comparison between regional and tertiary hospitals, showing significantly higher resistance levels in tertiary facilities. Error bars represent 95% confidence intervals, with sample size (n) labeled above each mean point.

## 4.0 Discussion

This study investigated the prevalence, distribution, and antibiotic resistance profiles of ESKAPEEc pathogens causing BSIs in KwaZulu-Natal, South Africa. Our results confirmed ESKAPE and *E. coli* as common causative bacteria in BSIs in children and adults and highlighted their diverse antibiotic susceptibility profiles.

One hundred and fifty-nine (159, 85.9%) isolates were identified as ESKAPE and *E. coli*. *K. pneumoniae* (28.9%) and *S. aureus* (28.3%) were the leading isolated pathogens, followed by *E. coli* (15.7%). The dominance of *K. pneumoniae* and *S. aureus* in the current study corroborates the findings of Ismail and Perovic (2018), who studied antimicrobial susceptibility profiles of ESKAPE pathogens isolated from patients with bacteraemia in South Africa from 2016-2018. The occurrence of *E. coli* (15.7%) in this study was slightly lower than 19% and 36% from the public and private hospitals reported in another South African study that analyzed antimicrobial susceptibility patterns of ESKAPE organisms isolated from patients with bacteraemia between 2016-2017. Their results showed that *E. coli* was the most common organism, followed by *K. pneumoniae* (Ismail et al., 2019).

The overall antibiotic susceptibility profiles showed that more than two-thirds of the *Enterobacterales* and *non-Enterobacterales* isolates (≥50%) were resistant to antibiotics in the Access category (ampicillin, amoxicillin-clavulanic acid, gentamycin) and Watch category (cephalosporins, piperacillin-tazobactam, ciprofloxacin), similar to that reported by Ismail and Perovic, (2018). Choonara et al., (2022), also observed resistance to cephalosporins, ciprofloxacin, gentamycin and ttrimethoprim-sulfamethoxazole in a study that was done in Malawi which investigated the antimicrobial susceptibility profiles of ESKAPEEc isolates from clinical samples including blood. However, their analysis differs from ours because their sample size was larger and included urine, blood, and pus specimens. Their study also identified *Proteus mirabilis* and *Salmonella* spp., which were included for further analysis. The majority (≥82%) of isolates in this study displayed high susceptibility to antibiotics in reserve category such as tigecycline. The high susceptibilities recorded against tigecycline could be due to the reserved use of tigecycline, mainly for the treatment of difficult to treat bacterial infections caused by MDR bacteria (Korczak *et al*., 2024). Therefore, high susceptibility to tigecycline suggest that they may still be relied upon in treating infections caused by *Enterobacterales* and *non-Enterobacterales*, although resistance has been documented (Jean *et al*., 2022).

In this study, 90% of *A. baumannii* and 27.2% of *K. pneumoniae* isolates were resistant to imipenem and meropenem (Table 1). The high resistance to carbapenems observed in this study (90%) among *A. baumannii* isolates is similar to that reported by Lowe et al., (2022) where more than 80% resistance to meropenem and imipenem was recorded. This is an important finding as treatment options for carbapenem resistant *Enterobacterales* (CRE) infections are generally limited to the use of Reserve antibiotics such as tigecycline and colistin, even though resistance against these antibiotics too is progressively increasing (Wang et al., 2018). In this study, we observed a relatively low levels of tigecycline resistance in *K. pneumoniae* (8.7%), *E. coli* (8.0%), *E. cloacae* (11.1%), and *A. baumannii* (9.1%) (Table 1). However, resistance to tigecycline should be closely monitored as it is reserved for the treatment of infections caused by MDR pathogens. The *E. cloacae* displayed high susceptibility to tigecycline, indicating that it may still be relied upon for effective treatment of infections caused by *E. cloacae* isolates

In total 44 (97.8%) *S. aureus*, displayed multidrug resistance, with isolates displaying high resistance to penicillin (93.3%), rifampicin (62.3%), tetracycline (60%), moxifloxacin (60.0%), ciprofloxacin (57.8%). This contrasts with a South African study investigating patients with *S. aureus* bacteremia (SAB), which reported high susceptibility to antibiotics tested except for penicillin (Steinhaus *et al*., 2018). Resistance to antibiotics in the Watch category like rifampicin (62.2%), moxifloxacin (60.0%), ciprofloxacin (57.8%), is of particular concern, and indicates a potential emerging resistance. The 28.9% MRSA prevalence reported in this study is similar to 27% reported in a South African study that investigated molecular epidemiology and virulence characteristics of *S. aureus* isolates from bacteraemic patients at Tygerberg Hospital, Cape Town (Abdulgader *et al*., 2020). However, *S. aureus* isolates in this study exhibited high levels of sensitivity to Access antibiotics including chloramphenicol (82.2%), clindamycin (75.6%), Watch antibiotics such as vancomycin (100%), and reserve antibiotics such as tigecycline (93.3%), quinupristin-dalfopristin (93.3%), and linezolid (88.9%). Moodley and Perovic, (2017) also observed high susceptibilities to vancomycin, linezolid, chloramphenicol, and clindamycin which is comparable with the findings of this study. The high susceptibility to linezolid, and vancomycin indicates that they may still be relied upon in treating infections caused by *S. aureus*.

The *E. faecium* (100%) isolates in this study isolates were MDR and displayed resistance to ampicillin (80.0%), erythromycin (67%), streptomycin (66.7%), tetracycline (67%), imipenem (53.3%) and ciprofloxacin (53.3%). However, the *E. faecium* isolates displayed high susceptibility against antibiotics in Watch category such as vancomycin (93.3%), and teicoplanin (60%). Furthermore, *E. faecium* isolates displayed high susceptibility against reserve antibiotics such as quinupristin-dalfopristin (93.3%), tigecycline (93.3%), and linezolid (80%), implying that the drugs can still be relied upon for treatment of infections caused by MDR pathogens. Similarly, high susceptibility of *E. faecium* isolates was observed against linezolid, vancomycin, and teicoplanin in another South African study that determined the prevalence and AMR patterns of enterococci, including *E. faecium* isolated from blood cultures at the South African public hospitals from January 2016 to December 2020 (Mogokotleng *et al*., 2023). Most (94.9%) isolates displayed a multidrug resistance phenotype, with 151 resistance patterns (antibiograms) (Table S3). The assortment of 157 antibiograms observed in MDR isolates indicates a wide diversity of resistant phenotypes in different bacteria. The varying resistance patterns observed in all isolates, including *A. baumannii* isolates may limits treatment options (Vivas *et al*., 2019). Most of our study isolates were primarily isolated from the paediatric outpatient department (POPD), intensive care unit (ICU), medical wards, surgical wards, burn units, and the orthopedic outpatient department (OOPD), with most isolates from ICU. The distribution of pathogens in this study concurs with a review conducted by Pons and Ruiz, (2019), who stated that ESKAPEEc pathogens are frequently isolated in ICUs, with high levels of resistance to antibiotics commonly used in ICU. Most patients in this study (60.4%) were less than a year old. This is significant since neonates and infants are at high risk of infection because of their underdeveloped immune systems making them vulnerable infections especially in the ICU (Habyarimana *et al*., 2021). Neonatal sepsis is commonly associated with antibiotic resistance and is recognised as a global concern because of limited access to effective antimicrobials warranting a search for new antimicrobials (Li et al., 2020; Saharman et al., 2021).

The MARIs observed in this study were largely high, with more than, 148 (93.1%) of isolates having MARI of ≥0.20 (Figure 1) indicating high selective pressure and that isolates originate from a high-risk source where antibiotics are regularly used (Joseph *et al*., 2017). *A. baumannii* (45.5%), and *K. pneumoniae* (32.6%) isolates were the leading pathogens isolated from ICU participants. Most isolates from ICU participants showed multidrug resistance phenotype, with isolates displaying high resistance to commonly used antibiotics in the Access category such as ampicillin, amoxicillin clavulanic acid, and those in the Watch category including piperacillin-tazobactam, cephalosporins, and carbapenems (Table 1). Similar high resistance patterns have been observed against these antibiotics in South Africa (Perovic *et al*., 2022), and India (Saxena *et al*., 2019). The MARI values for both *A. baumannii* and *K. pneumoniae* isolates from ICU was higher than ≥0.4 indicating isolates that these isolates were from environments where antibiotics are frequently used, as would be expected in ICU environments (Johnston *et al*., 2018). The observed variation in MARI values across clinical settings underscores the significant influence of healthcare environments on antimicrobial resistance (AMR) patterns. Although the overall difference in MARI across ward groups was statistically significant, post hoc analysis indicated that the only meaningful pairwise difference occurred between Intensive Care Units (ICUs) and Outpatient/Clinic-Based Services. This observation aligns with existing evidence that ICUs are high-risk zones for AMR due to the routine use of broad-spectrum antibiotics, frequent empirical treatment, invasive procedures, and prolonged patient stays all of which contribute to intensified antimicrobial selection pressure (Tacconelli et al., 2018; Vincent et al., 2009). In contrast, outpatient services are characterized by shorter patient interactions and lower acuity cases, resulting in comparatively reduced resistance levels (WHO, 2017). The significantly higher MARI values observed in tertiary hospitals compared to regional facilities further highlight the elevated risk of resistance emergence in referral-level institutions. Tertiary hospitals often manage more complex cases and serve as referral centers for critically ill patients, leading to increased antibiotic exposure and a higher prevalence of multidrug-resistant organisms (Laxminarayan et al., 2013). These findings emphasize the need for context-specific antimicrobial stewardship (AMS) interventions, particularly targeting high-risk environments such as ICUs and tertiary healthcare settings.

Overall, the alarming rates of antimicrobial resistance (AMR) identified in this study reflect a complex interplay of microbiological and systemic healthcare factors within the public sector in KwaZulu-Natal. These findings are particularly concerning in the context of empiric therapy for bloodstream infections, where timely and effective treatment is critical. The detection of phenotypic resistance to carbapenems among *Klebsiella pneumoniae* and *Acinetobacter baumannii* isolates suggests the presence of carbapenemase-producing organisms, likely mediated by resistance genes such as *bla_NDM_*, *bla_OXA-23_*, or *bla_OXA-48_*, which have been increasingly reported in South African tertiary hospitals (Perovic et al., 2020; Lowe et al., 2022, Marais et al., 2024). Similarly, the identification of the *mecA* gene in *Staphylococcus aureus* highlights the burden of methicillin-resistant strains (MRSA), further restricting treatment options and increasing reliance on last-line agents such as linezolid and vancomycin (Shuping & Jooste, 2017). These resistance mechanisms exacerbate clinical outcomes, contributing to prolonged hospital stays, increased healthcare costs, and heightened mortality risk.

Several factors within the local healthcare setting likely contribute to the high prevalence of resistance observed. Unregulated antibiotic prescribing especially for agents such as third-generation cephalosporins and amoxicillin-clavulanate remains a major driver of resistance, as previously documented in the region (Ramsamy et al., 2018). Compounding this issue is the limited implementation of robust antimicrobial stewardship (AMS) programs. Many public hospitals face shortages of specialized personnel, including clinical microbiologists and infectious disease practitioners, which hinders effective oversight and enforcement. A recent AMS situational analysis across KwaZulu-Natal public sector facilities confirmed critical gaps in infrastructure and stewardship practices (Chetty et al., 2022). Additional contributors may include widespread empirical antibiotic use, restricted access to rapid diagnostics, and prolonged ICU admissions, all of which increase the risk of horizontal gene transfer in high-risk hospital wards. These findings underscore the need for urgent, coordinated action. Strengthening antimicrobial stewardship programs across healthcare institutions should be prioritized through strategies such as formulary restrictions for Watch and Reserve antibiotics, regular audit-and-feedback mechanisms, and strict adherence to national treatment guidelines (Chetty et al., 2022). Investment in diagnostic microbiology, including molecular methods to detect key resistance genes, will facilitate timely, targeted therapy. Moreover, integrating IPC (Infection Prevention and Control) measures with real-time AMR surveillance will be essential for minimizing nosocomial transmission and guiding evidence-based interventions (Naidoo et al., 2019).

The limitations of this study include the absence of epidemiological data such as prior antibiotic exposure, treatment history, and clinical parameters which restricted our ability to assess risk factors for antimicrobial resistance. Additionally, the study was limited to culture-confirmed bloodstream infection (BSI) cases recorded in hospital databases. As a result, patients with less severe infections or those managed in outpatient settings may have been excluded, potentially limiting the generalizability of the findings to the broader patient population. Future studies should aim to incorporate larger, more comprehensive datasets including clinical and treatment information to enable more risk factor analysis and improve the external validity of resistance surveillance.

## Conclusion

This study demonstrates a high prevalence of multidrug resistance (MDR) among ESKAPEEc pathogens isolated from bloodstream infections, along with considerable variability in their resistance profiles. Notably, widespread resistance to WHO Watch group antibiotics was observed, potentially necessitating increased reliance on Reserve category agents for effective treatment. These findings underscore the urgent need for strengthened antimicrobial stewardship and routine AMR surveillance, particularly within public hospital settings, to inform treatment strategies and curb the spread of resistant pathogens.

## Supplementary Materials

**Table S1**: Demographic characteristics of patients; **Table S2**: Oligonucleotide primers and cycling conditions for the detection of ARGs; **Table S3:** Multiple antibiotic resistance index (MARI) of ESKAPEEc isolates; **Table S4:** Multiple antibiotic-resistant phenotypes displayed by ESKAPE and *E. coli* isolates; **Table S5**: Detailed phenotypic profiles of S. aureus isolates; **Table S6:** Detailed phenotypic profiles of E. faecium isolates; **Table S7:** Multiple antibiotic resistant phenotypes displayed by ESKAPE and *E. coli* isolates. **Table S8: A.** Results of Shapiro–Wilk and Levene’s tests for normality and homogeneity of variances**. B.** One-way ANOVA summary statistics for MARI values across ward groups and facility levels**. C.** Tukey’s HSD post hoc pairwise comparisons of MARI values among ward groups. **Table S9: A.** Descriptive statistics of MARI values by ward group, including mean, standard deviation, sample size, and 95% confidence intervals. **B.** Descriptive statistics of MARI values by facility level (regional vs tertiary), including mean, standard deviation, sample size, and 95% confidence intervals.

## Data availability

The generated data used to support the findings of this study are included in the main article and its supplementary materials.

## Author Contributions

Co-conceptualization, B.A; S.Y.E; laboratory work, BA; data analysis, B.A; write-up BA; critical review, editing, and formatting of manuscript B.A., J, A, J.M, J.A, S.Y.E, A.L.K.A., co-supervision: S.Y.E, D.G.A, A.L.K..A, J.M, and; funding acquisition, S.Y. E. All authors have read and agreed to the published version of the manuscript.

## Author approval and Data integrity

All authors have read and approved the final version of the manuscript. The corresponding author, Bakoena A. Hetsa, had full access to all of the data in this study and takes complete responsibility for the integrity of the data and the accuracy of the data analysis.

## Acknowledgements

The authors thank the National Health Laboratory Services (NHLS), for providing isolates and technical assistance.

## Funding

This work was funded by the South African Research Chairs Initiative of the Department of Science and Technology and National Research Foundation of South Africa (Grant No. 98342), the South African Medical Research Council (SAMRC), and UK Medical Research Council Newton Fund and the SAMRC Self-Initiated Research Grant, all awarded to S.Y.E. The funders had no role in the study’s design, in the collection, analyses, or interpretation of data, in the writing of the manuscript, or in the decision to publish the results. Any opinion, finding, and conclusion or recommendation expressed in this material is that of the authors.

## Conflicts of Interest

S.Y.E. is a chairperson of the Global Respiratory Partnership and a member of the Global Hygiene Council, both supported by unrestricted educational grants from Reckitt (Pty.) Ltd, UK. All other authors have no competing interests.

## Transparency Statement

Bakoena A. Hetsa affirms that this manuscript is an honest, accurate, and transparent account of the study being reported; that no important aspects have been omitted; and that any discrepancies from the study as planned have been explained.

## Notes

### Competing Interest Statement

The authors have declared no competing interest.

## References

1. Abdulgader SM, et al. The association between pathogen factors and clinical outcomes in patients with *Staphylococcus aureus* bacteraemia in a tertiary hospital, Cape Town. Int J Infect Dis. 2020;91:111–118. doi:10.1016/j.ijid.2019.11.032.

2. Amoako DG, et al. Genomic analysis of methicillin-resistant *Staphylococcus aureus* isolated from poultry and occupational farm workers in uMgungundlovu District, South Africa. Sci Total Environ. 2019;670:704–716. doi:10.1016/j.scitotenv.2019.03.110.

3. Chetty S, Reddy M, Ramsamy Y, Dlamini VC, Reddy-Naidoo R, Essack SY. Antimicrobial stewardship in public-sector hospitals in KwaZulu-Natal, South Africa. Antibiotics (Basel). 2022;11(7):881. doi:10.3390/antibiotics11070881.

4. Chen L, et al. Multiplex real-time PCR assay for detection and classification of *Klebsiella pneumoniae* carbapenemase gene (*blaKPC*) variants. J Clin Microbiol. 2011;49(2):579–585. doi:10.1128/JCM.01588-10.

5. Choonara FE, et al. Antimicrobial susceptibility profiles of clinically important bacterial pathogens at the Kamuzu Central Hospital in Lilongwe, Malawi. Malawi Med J. 2022;34(1):9–16.

6. CLSI. Performance standards for antimicrobial susceptibility testing. 27th ed. CLSI supplement M100. Wayne, PA: Clinical and Laboratory Standards Institute; 2017. Available at: www.clsi.org.

7. Crichton H, et al. Neonatal and paediatric bloodstream infections: Pathogens, antimicrobial resistance patterns and prescribing practice at Khayelitsha District Hospital, Cape Town, South Africa. S Afr Med J. 2018;108(2):99–104. doi:10.7196/SAMJ.2018.v108i2.12601.

8. De Angelis G, Posteraro B, De Carolis E, et al. Incidence and antimicrobial resistance trends in bloodstream infections caused by ESKAPE and *Escherichia coli* at a large teaching hospital in Rome, a 9-year analysis (2007–2015). Eur J Clin Microbiol Infect Dis. 2018;37(9):1627–1636. doi:10.1007/s10096-018-3292-9.

9. De Socio GV, Bartoletti M, Righi E, et al. Measurement and prediction of antimicrobial resistance in bloodstream infections by ESKAPE pathogens and *Escherichia coli*. J Glob Antimicrob Resist. 2019;19:154–160. doi:10.1016/j.jgar.2019.05.013.

10. EUCAST European Committee on Antimicrobial Susceptibility Testing. Breakpoint tables for interpretation of MICs and zone diameters. Version 8. 2017. Available at: https://www.eucast.org/clinical_breakpoints.

11. Gezmu AM, et al. Laboratory-confirmed bloodstream infections in two large neonatal units in sub-Saharan Africa. Int J Infect Dis. 2021;103:201–207. doi:10.1016/j.ijid.2020.11.169.

12. Habyarimana T, et al. Bacteriological profile and antimicrobial susceptibility patterns of bloodstream infection at Kigali University Teaching Hospital. Infect Drug Resist. 2021;14:699–707. doi:10.2147/IDR.S299520.

13. Holt KE, et al. Genomic analysis of diversity, population structure, virulence, and antimicrobial resistance in *Klebsiella pneumoniae*, an urgent threat to public health. Proc Natl Acad Sci U S A. 2015;112(27):E3574–E3581. doi:10.1073/pnas.1501049112.

14. Ismail H, Perovic O. Overview of antimicrobial susceptibility patterns of ESKAPE organisms isolated from patients with bacteraemia in South Africa, 2016–2018. NICD Surveillance Bulletin. 2018;18(1):2016–2018.

15. Ismail H, et al. Surveillance and comparison of antimicrobial susceptibility patterns of ESKAPE organisms isolated from patients with bacteraemia in South Africa, 2016– 2017. S Afr Med J. 2019;109(12):934–940. doi:10.7196/SAMJ.2019.v109i12.14079.

16. Jean S-S, Harnod D, Hsueh P-R. Global threat of carbapenem-resistant gram-negative bacteria. Front Cell Infect Microbiol. 2022;12:823684. doi:10.3389/fcimb.2022.823684.

17. Johnston D, et al. Usage of antibiotics in the intensive care units of an academic tertiary-level hospital. S Afr J Infect Dis. 2018;33(4):106–113. doi:10.1080/23120053.2018.1482645.

18. Joseph AA, et al. Multiple antibiotic resistance index of *Escherichia coli* isolates in a tertiary hospital in South-West Nigeria. Med J Zambia. 2017;44(4):225–232.

19. Korczak L, et al. Molecular mechanisms of tigecycline-resistance among Enterobacterales. Front Cell Infect Microbiol. 2024;14:1289396. doi:10.3389/fcimb.2024.1289396.

20. Krumperman PH. Multiple antibiotic resistance indexing of *Escherichia coli* to identify high-risk sources of fecal contamination of foods. Appl Environ Microbiol. 1983;46(1):165–170.

21. Laxminarayan R, et al. Antibiotic resistance—the need for global solutions. Lancet Infect Dis. 2013;13(12):1057–1098.

22. Li G, et al. Towards understanding global patterns of antimicrobial use and resistance in neonatal sepsis: Insights from the NeoAMR network. Arch Dis Child. 2020;105(1):26–31. doi:10.1136/archdischild-2019-316816.

23. Lowe M, Shuping L, Perovic O. Carbapenem-resistant Enterobacterales in patients with bacteraemia at tertiary academic hospitals in South Africa, 2019–2020: An update. S Afr Med J. 2022;112(8):542–552. doi:10.7196/SAMJ.2022.v112i8.16351.

24. Marais G, et al. Carbapenem-resistant *Klebsiella pneumoniae* among hospitalized patients in Cape Town, South Africa: molecular epidemiology and characterization. JAC Antimicrob Resist. 2024;6(2):dlae050. doi:10.1093/jacamr/dlae050.

25. Martinez RM, Wolk DM. Bloodstream infections. In: Diagnostic Microbiology of the Immunocompromised Host. 2016:653–689. doi:10.1128/microbiolspec.DMIH2-0031-2016.

26. Mogokotleng R, et al. A retrospective analysis of culture-confirmed enterococci bloodstream infections in South Africa, 2016–2020: A cross-sectional study. Trop Med Infect Dis. 2023;8(1):19. doi:10.3390/tropicalmed8010019.

27. Molechan C, et al. Molecular epidemiology of antibiotic-resistant *Enterococcus* spp. from the farm-to-fork continuum in intensive poultry production in KwaZulu-Natal, South Africa. Sci Total Environ. 2019;692:868–878. doi:10.1016/j.scitotenv.2019.07.324.

28. Moodley AS, Perovic O. Characterisation of Staphylococcus aureus bloodstream isolates from Gauteng and Western Cape Provinces, South Africa, 2016 and 2017. NICD Public Health Surveillance Bulletin. 2017;16(2):99–107.

29. Mpinda-Joseph P, et al. Healthcare-associated infections including neonatal bloodstream infections in a leading tertiary hospital in Botswana. Hosp Pract. 2019;47(4):203–210. doi:10.1080/21548331.2019.1650608.

30. Musicha P, et al. Trends in antimicrobial resistance in bloodstream infection isolates at a large urban hospital in Malawi (1998–2016): A surveillance study. Lancet Infect Dis. 2017;17(10):1042–1052. doi:10.1016/S1473-3099(17)30394-8.

31. Naidoo R, Naicker P, Lalloo U, Essack SY. A review of IPC and AMR strategies in South Africa: Implementation challenges and opportunities. Antibiotics. 2019;8(3):100. doi:10.3390/antibiotics8030100.

32. Perovic O, et al. Carbapenem-resistant Enterobacteriaceae in patients with bacteraemia at tertiary hospitals in South Africa, 2015 to 2018. Eur J Clin Microbiol Infect Dis. 2020;39(7):1287–1294. doi:10.1007/s10096-020-03845-4.

33. Pons MJ, Ruiz J. Current trends in epidemiology and antimicrobial resistance in intensive care units. J Emerg Crit Care Med. 2019;3:5. doi:10.21037/jeccm.2019.01.05.

34. Ramsamy Y, Essack SY, Sartorius B, Patel M, Mlisana KP. Antibiotic resistance trends of ESKAPE pathogens in KwaZulu-Natal, South Africa: A five-year retrospective analysis. Afr J Lab Med. 2018;7(2):887. doi:10.4102/ajlm.v7i2.887.

35. Rudd KE, et al. Global, regional, and national sepsis incidence and mortality, 1990– 2017: Analysis for the Global Burden of Disease Study. Lancet. 2020;395(10219):200–211. doi:10.1016/S0140-6736(19)32989-7.

36. Saharman YR, Karuniawati A, Severin JA, Verbrugh HA. Infections and antimicrobial resistance in intensive care units in lower-middle income countries: A scoping review. Antimicrob Resist Infect Control. 2021;10(1):1–19. doi:10.1186/s13756-020-00871-x.

37. Sands K, Carvalho MJ, Portal E, et al. Characterization of antimicrobial-resistant gram-negative bacteria that cause neonatal sepsis in seven low-and middle-income countries. Nat Microbiol. 2021;6(4):512–523. doi:10.1038/s41564-021-00870-7.

38. Santajit S, Indrawattana N. Mechanisms of antimicrobial resistance in ESKAPE pathogens. Biomed Res Int. 2016;2016:2475067. doi:10.1155/2016/2475067.

39. Saxena S, et al. Antimicrobial consumption and bacterial resistance pattern in patients admitted in ICU at a tertiary care center. J Infect Public Health. 2019;12(5):695–699. doi:10.1016/j.jiph.2019.03.014.

40. Shuping LL, Kuonza L, Musekiwa A, Iyaloo S, Perovic O. Hospital-associated methicillin-resistant *Staphylococcus aureus*: A cross-sectional analysis of risk factors in South African tertiary public hospitals. PLoS One. 2017;12(11):e0188216. doi:10.1371/journal.pone.0188216.

41. Steinhaus N, et al. The management and outcomes of *Staphylococcus aureus* bacteraemia at a South African referral hospital: A prospective observational study. Int J Infect Dis. 2018;73:78–84. doi:10.1016/j.ijid.2018.06.004.

42. Tacconelli E, et al. Discovery, research, and development of new antibiotics: The WHO priority list of antibiotic-resistant bacteria and tuberculosis. Lancet Infect Dis. 2018;18(3):318–327.

43. Tian L, Zhang Z, Sun Z. Antimicrobial resistance trends in bloodstream infections at a large teaching hospital in China: A 20-year surveillance study (1998–2017). Antimicrob Resist Infect Control. 2019;8(1):1–8. doi:10.1186/s13756-019-0545-z.

44. Vincent JL, et al. International study of the prevalence and outcomes of infection in intensive care units. JAMA. 2009;302(21):2323–2329.

45. Vivas R, et al. Multidrug-resistant bacteria and alternative methods to control them: An overview. Microb Drug Resist. 2019;25(6):890–908. doi:10.1089/mdr.2018.0319.

46. Wang Q, et al. Phenotypic and genotypic characterization of carbapenem-resistant Enterobacteriaceae: Data from a longitudinal large-scale CRE study in China (2012– 2016). Clin Infect Dis. 2018;67(2):196–205. doi:10.1093/cid/ciy660.

47. WHO. Antibacterial agents in clinical development: An analysis of the antibacterial clinical development pipeline. 2017. https://www.who.int/publications/i/item/antibacterial-agents-in-clinical-development

48. WHO. Bacterial priority pathogens list, bacterial pathogens of public health importance to guide research, development and strategies to prevent and control antimicrobial resistance. 2024. https://www.who.int/publications/i/item/9789240093461

