## Supplementary Materials for "Multidrug-Resistant ESKAPEEc Pathogens from Bloodstream Infections in South Africa: A Cross-Sectional Study Assessing Resistance to WHO AWaRe Antibiotics"

**Supplementary Table 1:** Oligonucleotide primers and cycling conditions for the detection of genes in this study

| Organism | Gene | Primer sequence (5′-3′) | PCR Program | Amplicon size (bp) | Reference |
|---|---|---|---|---|---|
| *E. faecium* | *sodA* | F-TACTGACAAACCATTCATGATG<br>R-AACTTCGTCACCAACGCGAAC | UNG 50s 98 °C, 50s 95 °C, 1min 55 °C, 1min 72°C | 112 | (Furlaneto-Maia et al., 2014) (Jackson et al., 2004) |
| *E. cloacae* | *hsp60* | F-GGTAGAAGAAGGCGTGGTTGC<br>R-ATGCATTCGGTGGTGATCATCAG | 5min 95°C, 30s 57°C, 1min 72°C | 341 | (Bakhshi et al., 2019) |
| *S. aureus* | *nuc* | F-ATGAACAACGTTCTGAAATTCTCTGCT<br>R- CTTGCGGCTGGCTTTTTCCAG | 5min 95°C, 30s 57°C, 1min 72°C | 270 | (Javid et al., 2018) |
| *S. aureus* | *mecA* | F- ACAGGTGAATTATTAGCACTTGTAAG<br>R- ATTGCTGTTAATATTTTTTGAGTTGAA | 5min 94°C, 55°C, 45s, 45s 72°C | 174 | (Asante et al., 2019) |
| *S. aureus* | *mecA* | F-′AACAGGTGAATTATTAGCACTTGTAAG3′<br>R-5′ATTGCTGTTAATATTTTTTGAGTTGAA3′ | UNG 5min 94 °C, 30s 94 °C, 30s, 62 °C, 45s, 72°C | | (Amoako et al., 2019) |
| *K. pneumoniae* | *khe* | F-CATCTGCCACACCTTTCTCA<br>R-CCGGGATTGAGCGGGTAATA | UNG 50s 98 °C, 10s 94 °C , 30s 55.8 °C, 1min 72°C | 400 | (Hartman et al., 2009) |
| *A. baumannii* | *sp4* F | F-CACGCCGTAAGAGTGCATTA<br>R-AACGGAGCTTGTCAGGGTTA | 5min 95°C, 30s 60°C, 30s 72°C | 490 | (Higgins et al., 2007) |
| *P. aeruginosa* | *oprl* F | F-ATGAACAACGTTCTGAAATTCTCTGCT<br>R-CTTGCGGCTGGCTTTTTCCAG | 5min 95°C, 30s 57°C, 1min 72°C | 249 | (Gholami et al., 2016) |
| *E. coli* | *uida* | F-AAAACGGCAAGAAAAAGCAG<br>R-ACGCGTGGTTAACAGTCTTGCG | UNG 50s 98 °C, 10s 95 °C, 30s 62 °C, 1min 72°C | 162 | (Godambe et al., 2017) |

**Supplementary Table 2:** Demographic characteristic characteristics of patients

| Isolate ID | Species | Sample | Facility Level | Ward Group | Department | Gender |
|---|---|---|---|---|---|---|
| K1 | *K. pneumoniae* | Blood | Regional | Intensive Care Units | Intensive care unit | F |
| K2 | *K. pneumoniae* | Blood | Regional | Outpatient/Clinic-Based Services | KMMC CLINIC | U |
| K3 | *K. pneumoniae* | Blood | Regional | Intensive Care Units | Intensive care unit | F |
| K4 | *K. pneumoniae* | Blood | Regional | Intensive Care Units | Intensive care unit | F |
| K5 | *K. pneumoniae* | Blood | Regional | General Inpatient Wards | Medical ward | F |
| K6 | *K. pneumoniae* | Blood | Regional | Surgical Wards | Surgical ward | M |
| K7 | *K. pneumoniae* | Blood | Tertiary | Emergency/Trauma Units | Casualty | F |
| K8 | *K. pneumoniae* | Blood | Tertiary | Intensive Care Units | Intensive care unit | F |
| K9 | *K. pneumoniae* | Blood | Regional | Specialist Wards | Burns ward | M |
| K10 | *K. pneumoniae* | Blood | Tertiary | Intensive Care Units | Intensive care unit | M |
| K12 | *K. pneumoniae* | Blood | Regional | Intensive Care Units | Intensive care unit | F |
| K13 | *K. pneumoniae* | Blood | Tertiary | Specialist Wards | Renal unit | F |
| K14 | *K. pneumoniae* | Blood | Regional | General Inpatient Wards | Medical ward | M |
| K15 | *K. pneumoniae* | Blood | Regional | Surgical Wards | Surgical ward | M |
| K21 | *K. pneumoniae* | Blood | Regional | Outpatient/Clinic-Based Services | KMMC CLINIC | M |
| K22 | *K. pneumoniae* | Blood | Tertiary | Intensive Care Units | Intensive care unit | M |
| K23 | *K. pneumoniae* | Blood | Regional | Surgical Wards | Surgical ward | F |
| K24 | *K. pneumoniae* | Blood | Tertiary | Intensive Care Units | Intensive care unit | M |
| K26 | *K. pneumoniae* | Blood | Regional | Surgical Wards | Surgical ward | M |
| K28 | *K. pneumoniae* | Blood | Tertiary | Specialist Wards | Renal unit | M |
| K29 | *K. pneumoniae* | Blood | Regional | Surgical Wards | Surgical ward | M |
| K30 | *K. pneumoniae* | Blood | Regional | Paediatric Units | Paediatric ward | M |
| K31 | *K. pneumoniae* | Blood | Regional | Emergency/Trauma Units | EMERG DEPT | F |
| K32 | *K. pneumoniae* | Blood | Regional | Emergency/Trauma Units | EMERG DEPT | F |
| K33 | *K. pneumoniae* | Blood | Regional | Paediatric Units | Paediatric ward | F |
| K34 | *K. pneumoniae* | Blood | Tertiary | Intensive Care Units | Intensive care unit | F |
| K35 | *K. pneumoniae* | Blood | Regional | Paediatric Units | Paediatric ward | M |
| K36 | *K. pneumoniae* | Blood | Tertiary | Intensive Care Units | Intensive care unit | F |
| K37 | *K. pneumoniae* | Blood | Regional | Paediatric Units | Paediatric ward | M |
| K38 | *K. pneumoniae* | Blood | Regional | Intensive Care Units | Surgical intensive care unit | F |
| K39 | *K. pneumoniae* | Blood | Regional | Intensive Care Units | Surgical intensive care unit | M |
| K40 | *K. pneumoniae* | Blood | Tertiary | Intensive Care Units | Intensive care unit | M |

| | | | | | | |
|---|---|---|---|---|---|---|
| K41 | *K. pneumoniae* | Blood | Tertiary | Specialist Wards | Renal unit | F |
| K42 | *K. pneumoniae* | Blood | Regional | Paediatric Units | Paediatric ward | M |
| K43 | *K. pneumoniae* | Blood | Regional | Paediatric Units | Paediatric ward | M |
| K44 | *K. pneumoniae* | Blood | Regional | Paediatric Units | Paediatric ward | F |
| K45 | *K. pneumoniae* | Blood | Regional | General Inpatient Wards | Medical ward | F |
| K46 | *K. pneumoniae* | Blood | Regional | Paediatric Units | Paediatric ward | M |
| K47 | *K. pneumoniae* | Blood | Regional | Intensive Care Units | Intensive care unit | F |
| K48 | *K. pneumoniae* | Blood | Tertiary | Intensive Care Units | Intensive care unit | U |
| K49 | *K. pneumoniae* | Blood | Regional | General Inpatient Wards | Medical ward | M |
| K50 | *K. pneumoniae* | Blood | Regional | Intensive Care Units | Intensive care unit | F |
| K51 | *K. pneumoniae* | Blood | Regional | Specialist Wards | Burns ward | M |
| K52 | *K. pneumoniae* | Blood | Regional | General Inpatient Wards | Medical ward | F |
| K53 | *K. pneumoniae* | Blood | Tertiary | Intensive Care Units | Intensive care unit | U |
| E1 | *E. coli* | Blood | Regional | General Inpatient Wards | Medical ward | M |
| E2 | *E. coli* | Blood | Regional | General Inpatient Wards | Medical ward | U |
| E3 | *E. coli* | Blood | Tertiary | Emergency/Trauma Units | Casualty | F |
| E4 | *E. coli* | Blood | Regional | General Inpatient Wards | Medical ward | F |
| E5 | *E. coli* | Blood | Regional | Intensive Care Units | Intensive care unit | U |
| E6 | *E. coli* | Blood | Regional | Intensive Care Units | Intensive care unit | M |
| E7 | *E. coli* | Blood | Regional | Emergency/Trauma Units | Emergency department | F |
| E8 | *E. coli* | Blood | Regional | General Inpatient Wards | Medical ward | F |
| E10 | *E. coli* | Blood | Regional | Intensive Care Units | Surgical intensive care unit | M |
| E21 | *E. coli* | Blood | Tertiary | Surgical Wards | Surgical ward | F |
| E13 | *E. coli* | Blood | Regional | Emergency/Trauma Units | Emergency department | M |
| E16 | *E. coli* | Blood | Tertiary | Emergency/Trauma Units | Casualty | M |
| E17 | *E. coli* | Blood | Regional | Intensive Care Units | Surgical intensive care unit | M |
| E19 | *E. coli* | Blood | Regional | Intensive Care Units | Surgical intensive care unit | M |
| E26 | *E. coli* | Blood | Regional | Surgical Wards | Surgical ward | F |
| E30 | *E. coli* | Blood | Regional | Surgical Wards | Surgical ward | M |
| E12 | *E. coli* | Blood | Tertiary | Intensive Care Units | intensive care unit | M |
| E20 | *E. coli* | Blood | Tertiary | Intensive Care Units | intensive care unit | M |
| E28 | *E. coli* | Blood | Regional | Paediatric Units | Paediatric ward | M |
| E27 | *E. coli* | Blood | Regional | Intensive Care Units | Intensive care unit | F |

| | | | | | | |
|---|---|---|---|---|---|---|
| E24 | *E. coli* | Blood | Regional | Intensive Care Units | Intensive care unit | F |
| E11 | *E. coli* | Blood | Regional | Paediatric Units | Paediatric ward | F |
| E14 | *E. coli* | Blood | Regional | Outpatient/Clinic-Based Services | Paediatric outpatient | M |
| E18 | *E. coli* | Blood | Regional | Paediatric Units | Paediatric ward | F |
| E25 | *E. coli* | Blood | Regional | Intensive Care Units | Intensive care unit | M |
| EC3 | *E. cloacae* | Blood | Regional | Intensive Care Units | Surgical intensive care unit | M |
| EC10 | *E. cloacae* | Blood | Regional | Intensive Care Units | Surgical intensive care unit | F |
| EC9 | *E. cloacae* | Blood | Tertiary | Emergency/Trauma Units | Casualty | M |
| EA1 | *E. cloacae* | Blood | Regional | Intensive Care Units | intensive care unit | M |
| EC7 | *E. cloacae* | Blood | Tertiary | Intensive Care Units | Surgical intensive care unit | M |
| EC6 | *E. cloacae* | Blood | Tertiary | Specialist Wards | Dermatology clinic | F |
| EC8 | *E. cloacae* | Blood | Regional | Paediatric Units | Paediatric ward | F |
| EC5 | *E. cloacae* | Blood | Regional | Paediatric Units | Paediatric ward | M |
| EC2 | *E. cloacae* | Blood | Regional | Paediatric Units | Paediatric ward | F |
| A5 | *A. baumanii* | Blood | Regional | Surgical Wards | Surgical ward | M |
| A2 | *A. baumanii* | Blood | Tertiary | Surgical Wards | Surgical ward | F |
| A10 | *A. baumanii* | Blood | Regional | General Inpatient Wards | Medical ward | F |
| A12 | *A. baumanii* | Blood | Tertiary | Intensive Care Units | Intensive care unit | F |
| A3 | *A. baumanii* | Blood | Regional | Surgical Wards | Surgical ward | F |
| A6 | *A. baumanii* | Blood | Tertiary | Intensive Care Units | intensive care unit | U |
| A8 | *A. baumanii* | Blood | Tertiary | Intensive Care Units | intensive care unit | F |
| A11 | *A. baumanii* | Blood | Tertiary | Intensive Care Units | intensive care unit | F |
| A9 | *A. baumanii* | Blood | Regional | General Inpatient Wards | Paediatric outpatient | U |
| A1 | *A. baumanii* | Blood | Tertiary | Outpatient/Clinic-Based Services | Paediatric outpatient | M |
| A4 | *A. baumanii* | Blood | Regional | Intensive Care Units | intensive care unit | M |
| P5 | *P. aeruginosa* | Blood | Regional | Specialist Wards | Burns ward | M |
| P6 | *P. aeruginosa* | Blood | Regional | Specialist Wards | Burns ward | U |
| P2 | *P. aeruginosa* | Blood | Regional | Surgical Wards | Surgical ward | M |
| P7 | *P. aeruginosa* | Blood | Regional | Specialist Wards | Burns ward | M |
| P1 | *P. aeruginosa* | Blood | Regional | Surgical Wards | Surgical ward | M |
| P3 | *P. aeruginosa* | Blood | Regional | Outpatient/Clinic-Based Services | Paediatric outpatient | M |
| P8 | *P. aeruginosa* | Blood | Regional | Intensive Care Units | Intensive care unit | M |
| P4 | *P. aeruginosa* | Blood | Regional | Specialist Wards | Burns ward | M |
| S2 | *S. aureus* | Blood | Regional | General Inpatient Wards | Medical ward | F |
| S3 | *S. aureus* | Blood | Regional | Emergency/Trauma Units | Emergency department | F |

| | | | | | | |
|---|---|---|---|---|---|---|
| S11 | *S. aureus* | Blood | Regional | Surgical Wards | Surgical ward | F |
| S21 | *S. aureus* | Blood | Regional | Emergency/Trauma Units | Emergency department | M |
| S22 | *S. aureus* | Blood | Regional | General Inpatient Wards | Medical ward | F |
| S24 | *S. aureus* | Blood | Regional | Surgical Wards | Surgical ward | M |
| S25 | *S. aureus* | Blood | Regional | Surgical Wards | Surgical ward | M |
| S33 | *S. aureus* | Blood | Regional | Outpatient/Clinic-Based Services | Medical ward | F |
| S38 | *S. aureus* | Blood | Regional | General Inpatient Wards | Medical ward | F |
| S39 | *S. aureus* | Blood | Regional | Surgical Wards | Surgical ward | M |
| S41 | *S. aureus* | Blood | Regional | General Inpatient Wards | Medical ward | M |
| S42 | *S. aureus* | Blood | Regional | Outpatient/Clinic-Based Services | Orthopedic out-patient department | M |
| S43 | *S. aureus* | Blood | Regional | General Inpatient Wards | Medical ward | F |
| S49 | *S. aureus* | Blood | Regional | Emergency/Trauma Units | Emergency department | F |
| S18 | *S. aureus* | Blood | Regional | Outpatient/Clinic-Based Services | Extension ward | M |
| S30 | *S. aureus* | Blood | Regional | Specialist Wards | Burns ward | F |
| S31 | *S. aureus* | Blood | Regional | Specialist Wards | Burns ward | F |
| S13 | *S. aureus* | Blood | Regional | Paediatric Units | Paediatric ward | M |
| S46 | *S. aureus* | Blood | Regional | Outpatient/Clinic-Based Services | Extension ward | M |
| S6 | *S. aureus* | Blood | Tertiary | Intensive Care Units | Intensive care unit | M |
| S7 | *S. aureus* | Blood | Regional | Outpatient/Clinic-Based Services | Orthopedic out-patient department | F |
| S12 | *S. aureus* | Blood | Regional | Paediatric Units | Paediatric ward | F |
| S15 | *S. aureus* | Blood | Regional | Outpatient/Clinic-Based Services | Paediatric outpatient department | F |
| S19 | *S. aureus* | Blood | Regional | Outpatient/Clinic-Based Services | Paediatric outpatient department | M |
| S28 | *S. aureus* | Blood | Regional | Outpatient/Clinic-Based Services | Paediatric outpatient department | U |
| S29 | *S. aureus* | Blood | Regional | Paediatric Units | Paediatric ward | M |
| S32 | *S. aureus* | Blood | Regional | Intensive Care Units | Intensive care unit | M |
| S34 | *S. aureus* | Blood | Regional | Outpatient/Clinic-Based Services | Paediatric outpatient department | M |
| S40 | *S. aureus* | Blood | Regional | Outpatient/Clinic-Based Services | Paediatric outpatient department | M |
| S44 | *S. aureus* | Blood | Regional | Intensive Care Units | Intensive care unit | F |
| S45 | *S. aureus* | Blood | Regional | Outpatient/Clinic-Based Services | Orthopedic out-patient department | F |
| S48 | *S. aureus* | Blood | Tertiary | Intensive Care Units | Intensive care unit | U |
| S1 | *S. aureus* | Blood | Regional | Outpatient/Clinic-Based Services | Paediatric outpatient department | F |
| S4 | *S. aureus* | Blood | Regional | Outpatient/Clinic-Based Services | KMMC clinic | F |
| S5 | *S. aureus* | Blood | Regional | Paediatric Units | KMMC clinic | F |

| | | | | | | |
|---|---|---|---|---|---|---|
| S8 | *S. aureus* | Blood | Regional | Emergency/Trauma Units | Emergency department | M |
| S9 | *S. aureus* | Blood | Regional | Outpatient/Clinic-Based Services | Paediatric outpatient department | F |
| S10 | *S. aureus* | Blood | Regional | Outpatient/Clinic-Based Services | Paediatric outpatient department | M |
| S13 | *S. aureus* | Blood | Regional | Outpatient/Clinic-Based Services | Paediatric outpatient department | F |
| S14 | *S. aureus* | Blood | Regional | Outpatient/Clinic-Based Services | Orthopedic out-patient department | M |
| S16 | *S. aureus* | Blood | Regional | Outpatient/Clinic-Based Services | Paediatric outpatient department | F |
| S17 | *S. aureus* | Blood | Regional | Outpatient/Clinic-Based Services | Paediatric outpatient department | M |
| S20 | *S. aureus* | Blood | Regional | Surgical Wards | Surgical ward | M |
| S26 | *S. aureus* | Blood | Regional | Outpatient/Clinic-Based Services | Paediatric outpatient department | M |
| S37 | *S. aureus* | Blood | Regional | Paediatric Units | Paediatric ward | M |
| EF10 | *E. faecium* | Blood | Regional | Outpatient/Clinic-Based Services | Paediatric outpatient department | F |
| EF6 | *E. faecium* | Blood | Regional | Intensive Care Units | Intensive care unit | F |
| EF5 | *E. faecium* | Blood | Regional | Paediatric Units | Medical ward | F |
| EF7 | *E. faecium* | Blood | Tertiary | General Inpatient Wards | Medical ward | F |
| EF8 | *E. faecium* | Blood | Regional | Paediatric Units | Paediatric ward | F |
| EF9 | *E. faecium* | Blood | Regional | General Inpatient Wards | Medical ward | M |
| EF12 | *E. faecium* | Blood | Regional | Outpatient/Clinic-Based Services | Paediatric outpatient department | M |
| EF13 | *E. faecium* | Blood | Regional | Outpatient/Clinic-Based Services | Paediatric outpatient department | F |
| EF15 | *E. faecium* | Blood | Regional | Outpatient/Clinic-Based Services | Paediatric outpatient department | M |
| EF19 | *E. faecium* | Blood | Regional | Outpatient/Clinic-Based Services | KMM CLINIC | M |
| EF21 | *E. faecium* | Blood | Regional | Paediatric Units | Medical ward | M |
| EF23 | *E. faecium* | Blood | Regional | Intensive Care Units | Intensive care unit | M |
| EF24 | *E. faecium* | Blood | Tertiary | Surgical Wards | Surgical ward | F |
| EF25 | *E. faecium* | Blood | Regional | Outpatient/Clinic-Based Services | KMMC CLINIC | M |
| EF26 | *E. faecium* | Blood | Regional | Paediatric Units | Paediatric ward | M |

**Table S3:** Multiple antibiotic resistance index (MARI) of ESKAPEEc isolates

| MAR index | No of isolates (%) | | | | | | |
|---|---|---|---|---|---|---|---|
| | *E. faecium* | *S. aureus* | *K. pneumoniae* | *A. baumannii* | *P. aeruginosa* | *E. cloacae* | *E. coli* |
| 0.05 | 0 (0) | 0 (0) | 2 (4.3) | 0 (0) | 0 (0) | 0 (0) | 0 (0) |
| 0.10 | 0 (0) | 2 (4.4) | 0 (0) | 0 (0) | 0 (0) | 0 (0.0) | 0 (0) |
| 0.15 | 3 (20.0) | 0 (0) | 0 (0) | 0 (0) | 0 (0) | 1 (11.1) | 0 (0) |
| 0.20 | 0 (0) | 6 (13.3) | 0 (0) | 1 (9.1) | 0 (0) | 1 (11.1) | 1 (4.0) |
| 0.25 | 0 (0) | 3 (6.7) | 1 (2.2) | 0 (0) | 2 (25.0) | 2 (22.2) | 3 (12.0) |
| 0.30 | 2 (13.3) | 7 (15.6) | 2 (4.3) | 0 (0) | 1 (12.5) | 0 (0.0) | 1 (4.0) |
| 0.35 | 1(6.7) | 3 (6.7) | 5 (10.9) | 1 (9.1) | 3 (37.5) | 0 (0.0) | 0 (0) |
| 0.40 | 1(6.7) | 4 (8.9) | 1 (2.2) | 1 (9.1) | 0 (0) | 1 (11.1) | 2 (8.0) |
| 0.45 | 0 (0) | 5 (11.1) | 3 (6.5) | 1 (9.1) | 0 (0) | 2 (22.2) | 2 (8.0) |
| 0.50 | 2 (13.3) | 5 (11.1) | 2 (4.3) | 1 (9.1) | 0 (0) | 1 (11.1) | 3 (12.0) |
| 0.55 | 0 (0) | 1 (6.7) | 8 (17.4) | 1 (9.1) | 2 (25.0) | 0 (0) | 3 (12.0) |
| 0.60 | 2 (13.3) | 4 (8.9) | 9 (19.6) | 1 (9.1) | 0 (0) | 0 (0) | 3 (12.0) |
| 0.65 | 0 (0) | 3 (6.7) | 2 (4.3) | 1 (9.1) | 0 (0) | 0 (0) | 3 (12.0) |
| 0.70 | 0 (0) | 0 (0) | 3 (6.5) | 0 (0) | 0 (0) | 0 (0) | 1 (4.0) |
| 0.75 | 0 (0) | 2 (4.4) | 4 (8.7) | 0 (0) | 0 (0) | 2 (11.1) | 2 (8.0) |
| 0.80 | 0 (0) | 0 (0) | 4 (8.7) | 1 (9.1) | 0 (0) | 0 (0) | 0 (0) |
| 0.85 | 0 (0) | 0 (0) | 0 (0) | 1 (9.1) | 0 (0) | 0 (0) | 1 (4.0) |
| 0.90 | 0 (0) | 0 (0) | 0 (0) | 0 (0) | 0 (0) | 0 (0) | 0 (0) |
| 0.95 | 0 (0) | 0 (0) | 0 (0) | 0 (0) | 0 (0) | 0 (0) | 0 (0) |
| 1.0 | 0 (0) | 0 (0) | 0 (0) | 1 (9.1) | 0 (0) | 0 (0) | 0 (0) |

### Table S4: Detailed phenotypic profiles of Gram-negative isolates

| Isolate ID | Species | Ward type | Antibiotic resistance profile | | | | | | | | | | | | | | | | | | | |
|---|---|---|---|---|---|---|---|---|---|---|---|---|---|---|---|---|---|---|---|---|---|---|
| | | | AMP | AMC | TZP | CTX | CAZ | CRO | FEP | LEX | FOX | IMP | MEM | NAL | CIP | GEN | AMK | TGC | CHL | SXT | AZM | TET |
| E1 | *E. coli* | Med ward | S | S | I | S | I | I | I | R | R | I | I | I | S | I | I | S | S | R | S | S |
| E2 | *E. coli* | Med ward | R | R | S | I | R | R | R | R | S | S | S | S | S | I | I | S | S | R | S | S |
| E3 | *E. coli* | Casualty | R | R | S | R | R | R | R | R | S | I | S | R | R | R | R | I | S | R | R | R |
| E4 | *E. coli* | Med ward | R | R | I | R | R | R | R | R | S | I | I | R | R | R | R | I | S | R | R | R |
| E5 | *E. coli* | ICU | R | R | R | R | I | R | I | S | S | I | S | R | R | I | R | I | R | R | S | R |
| E6 | *E. coli* | ICU | R | R | R | R | R | R | R | R | S | I | S | I | S | S | R | S | S | R | S | R |
| E7 | *E. coli* | Emerg dpt | R | R | S | S | S | R | I | R | R | R | R | R | R | R | R | I | R | R | S | R |
| E8 | *E. coli* | Med ward | R | R | I | R | R | R | R | R | S | S | S | R | R | R | S | I | S | R | S | 1 |
| E10 | *E. coli* | SICU | R | R | I | I | I | R | I | R | R | R | R | R | R | S | S | S | R | R | R | R |
| E21 | *E. coli* | Surg ward | R | R | S | R | R | R | R | R | S | I | S | R | R | R | I | S | R | R | R | R |
| E13 | *E. coli* | Emerg dpt | R | R | R | R | R | R | R | R | 1 | I | S | R | R | R | R | I | S | R | R | R |
| E16 | *E. coli* | Casualty | R | R | I | R | R | R | R | R | S | I | S | R | R | R | R | S | S | R | R | R |
| E17 | *E. coli* | SICU | R | R | S | R | R | R | R | R | S | I | S | R | R | R | R | S | S | R | R | R |
| E19 | *E. coli* | SICU | R | R | S | S | S | I | I | S | S | S | S | R | R | R | I | S | 1 | R | R | R |
| E26 | *E. coli* | Surg ward | R | R | R | R | R | R | R | R | 1 | S | S | R | R | R | I | I | S | R | R | R |
| E30 | *E. coli* | Surg ward | R | R | R | R | R | R | R | R | 1 | S | S | R | R | R | I | I | S | R | R | R |
| E12 | *E. coli* | ICU | R | R | R | R | R | R | R | R | R | R | R | R | R | R | R | R | R | R | R | R |
| E20 | *E. coli* | ICU | R | R | R | R | R | R | R | R | S | S | S | R | R | R | R | I | R | R | R | R |
| E28 | *E. coli* | Paed ward | R | R | R | R | R | R | R | R | R | S | S | I | R | R | R | R | S | 1 | S | S |
| E27 | *E. coli* | ICU | R | R | R | R | R | R | R | R | R | S | S | I | R | R | I | R | S | 1 | S | S |
| E24 | *E. coli* | ICU | R | R | R | R | R | R | R | R | R | S | S | R | R | R | R | R | S | 1 | S | S |
| E11 | *E. coli* | Paed ward | R | R | S | I | I | S | I | S | R | S | S | I | R | I | I | I | S | R | R | R |
| E14 | *E. coli* | POPD | R | R | S | R | S | R | I | R | S | I | S | R | R | S | S | I | S | R | R | R |
| E18 | *E. coli* | Paed ward | R | R | S | R | R | R | R | R | R | I | S | R | R | R | I | I | R | R | R | R |
| E25 | *E. coli* | ICU | R | R | I | S | S | S | I | S | S | S | S | I | S | R | S | I | S | R | R | I |
| EC3 | *E. cloacae* | SICU | R | R | R | R | R | R | R | R | R | S | S | R | R | R | I | I | R | R | S | R |
| EC10 | *E. cloacae* | SICU | R | R | R | R | R | R | R | R | R | S | S | I | I | S | I | I | S | R | S | S |
| EC9 | *E. cloacae* | Casualty | R | R | I | R | R | R | R | R | R | S | S | I | I | S | S | I | S | R | S | S |
| EA1 | *E. cloacae* | ICU | R | S | S | S | S | R | I | S | S | S | S | I | R | S | I | I | S | S | S | S |
| EC7 | *E. cloacae* | SICU | R | R | I | R | R | I | I | R | R | S | S | S | R | S | I | R | S | S | S | S |
| EC6 | *E. cloacae* | Dermatolog y | R | R | S | R | S | 1 | R | R | R | S | S | S | R | S | I | I | S | S | R | S |
| EC8 | *E. cloacae* | Paed ward | R | R | I | I | I | R | I | R | R | S | S | S | S | S | S | I | R | S | S | S |

| | | | | | | | | | | | | | | | | | | | | | | |
|---|---|---|---|---|---|---|---|---|---|---|---|---|---|---|---|---|---|---|---|---|---|---|
| EC5 | *E. cloacae* | Paed ward | R | R | S | S | I | S | I | S | S | S | S | S | S | S | S | S | S | R | R | R |
| EC2 | *E. cloacae* | Paed ward | R | R | S | S | S | I | I | R | R | S | S | I | R | S | S | I | S | S | S | S |
| K1 | *K. pneumoniae* | ICU | NT | R | R | R | R | R | R | R | R | I | S | I | R | R | I | R | R | R | S | R |
| K2 | *K. pneumoniae* | KMMC CLINIC | NT | R | R | R | R | R | R | R | R | I | S | R | R | R | R | R | R | R | R | R |
| K3 | *K. pneumoniae* | ICU | NT | R | R | R | R | R | R | R | R | S | R | R | R | R | R | R | R | R | S | R |
| K4 | *K. pneumoniae* | ICU | NT | R | R | R | R | R | R | R | R | I | S | R | R | R | R | R | R | R | S | R |
| K5 | *K. pneumoniae* | Med ward | NT | R | R | R | R | R | R | R | R | R | S | R | R | R | R | R | R | R | S | I |
| K6 | *K. pneumoniae* | Surg ward | NT | R | R | R | R | R | R | R | R | R | R | R | R | R | R | R | R | R | R | R |
| K7 | *K. pneumoniae* | Casualty | NT | R | R | R | R | R | R | R | R | R | R | R | R | R | R | R | R | R | S | R |
| K8 | *K. pneumoniae* | ICU | NT | R | R | R | R | R | R | R | R | S | S | R | R | R | R | R | S | R | R | R |
| K9 | *K. pneumoniae* | Burns ward | NT | R | R | R | R | R | R | R | R | S | S | R | R | R | I | I | R | R | R | R |
| K10 | *K. pneumoniae* | ICU | NT | R | R | R | R | R | R | R | R | S | S | R | R | R | R | R | R | R | S | R |
| K12 | *K. pneumoniae* | ICU | NT | R | R | R | R | R | R | R | R | S | S | R | R | R | I | R | R | R | S | R |
| K13 | *K. pneumoniae* | Renal unit | NT | R | R | R | R | R | R | R | R | S | S | R | R | R | I | R | R | R | S | I |
| K14 | *K. pneumoniae* | Med ward | NT | R | R | R | R | R | R | R | R | S | S | R | R | R | I | R | R | R | S | R |
| K15 | *K. pneumoniae* | Surg ward | NT | R | R | I | R | R | R | R | R | S | S | I | S | R | I | R | R | S | R | R |
| K21 | *K. pneumoniae* | KMMC CLINIC | NT | R | R | R | R | R | R | R | R | S | S | R | R | R | S | I | S | R | S | S |
| K22 | *K. pneumoniae* | ICU | NT | R | R | S | R | R | R | S | R | S | R | I | R | S | S | R | S | R | S | R |
| K23 | *K. pneumoniae* | Surg ward | NT | R | R | R | R | R | R | R | R | S | S | R | R | R | R | R | R | R | S | R |
| K24 | *K. pneumoniae* | ICU | NT | R | R | R | R | R | R | R | R | R | R | R | R | R | I | R | S | R | S | R |
| K26 | *K. pneumoniae* | Surg ward | NT | R | R | S | I | S | R | S | R | S | S | I | R | S | I | R | S | S | S | S |
| K28 | *K. pneumoniae* | Renal unit | NT | R | R | R | R | R | R | R | R | S | S | R | R | R | S | I | R | R | S | S |
| K29 | *K. pneumoniae* | Surg ward | NT | R | R | R | R | R | R | R | R | S | 1 | R | R | R | R | R | R | R | R | R |
| K30 | *K. pneumoniae* | Paed ward | NT | R | R | R | R | R | R | R | R | S | S | R | R | R | R | R | S | R | S | S |
| K31 | *K. pneumoniae* | Emrg dept | NT | R | R | R | R | R | R | R | R | S | R | R | R | R | I | R | R | R | R | R |
| K32 | *K. pneumoniae* | Emrg dept | NT | R | R | R | R | R | R | R | R | R | R | R | R | R | I | R | R | R | R | R |
| K33 | *K. pneumoniae* | Paed ward | NT | R | R | R | R | R | R | R | R | S | S | R | R | R | R | R | R | R | R | R |
| K34 | *K. pneumoniae* | ICU | NT | R | R | R | R | R | R | R | R | S | R | R | R | R | S | R | S | R | S | R |
| K35 | *K. pneumoniae* | Paed ward | NT | R | R | R | R | R | R | R | R | S | R | R | R | R | I | I | S | R | S | R |
| K36 | *K. pneumoniae* | ICU | NT | R | R | R | R | R | R | R | R | S | R | R | R | R | R | R | R | R | R | R |
| K37 | *K. pneumoniae* | Paed ward | NT | R | R | R | R | R | R | R | R | S | S | R | R | R | S | R | S | R | S | R |
| K38 | *K. pneumoniae* | SICU | NT | R | R | I | S | R | R | R | R | S | R | R | R | R | I | R | S | S | S | I |
| K39 | *K. pneumoniae* | SICU | NT | R | R | R | R | R | R | R | S | S | S | R | R | R | I | I | R | R | S | R |
| K40 | *K. pneumoniae* | ICU | NT | R | R | R | R | R | R | R | S | S | S | R | R | R | I | I | R | R | S | R |
| K41 | *K. pneumoniae* | Renal unit | NT | R | R | R | R | R | R | R | S | S | S | I | I | R | S | I | S | S | S | S |
| K42 | *K. pneumoniae* | Paed ward | NT | R | R | R | R | R | R | R | R | R | R | R | R | R | I | R | R | R | R | R |

| | | | | | | | | | | | | | | | | | | | | | | |
|---|---|---|---|---|---|---|---|---|---|---|---|---|---|---|---|---|---|---|---|---|---|---|
| K43 | *K. pneumoniae* | Paed ward | NT | R | R | R | R | R | R | R | R | R | R | R | R | R | R | R | R | R | S | I |
| K44 | *K. pneumoniae* | Paed ward | NT | R | R | R | R | R | R | R | R | R | R | R | R | R | I | I | R | R | R | I |
| K45 | *K. pneumoniae* | Med ward | NT | S | S | S | S | 1 | I | S | S | S | S | S | S | S | S | S | S | S | S | S |
| K46 | *K. pneumoniae* | Paed ward | NT | R | R | R | R | R | S | R | S | S | S | R | R | R | R | I | S | 1 | S | S |
| K47 | *K. pneumoniae* | ICU | NT | S | S | S | S | S | I | S | S | S | S | S | S | S | S | S | S | S | S | S |
| K48 | *K. pneumoniae* | ICU | NT | R | R | R | R | R | R | R | R | S | S | R | R | R | I | I | R | R | R | R |
| K49 | *K. pneumoniae* | Med ward | NT | R | R | R | R | R | R | R | R | R | R | R | R | R | R | I | R | R | R | I |
| K50 | *K. pneumoniae* | ICU | NT | R | R | R | R | R | R | R | R | S | S | R | R | R | S | R | S | 1 | S | S |
| K51 | *K. pneumoniae* | Burns ward | NT | R | R | R | R | R | R | R | R | S | S | R | R | R | I | R | S | R | R | R |
| K52 | *K. pneumoniae* | Med ward | NT | R | R | R | R | R | R | R | R | R | R | R | R | R | R | R | R | R | R | I |
| K53 | *K. pneumoniae* | ICU | NT | R | I | R | R | R | R | R | S | S | S | R | R | R | S | I | R | R | S | R |
| A5 | *A. baumanii* | Surg ward | R | R | R | R | R | R | R | R | R | R | R | R | R | R | R | R | R | R | R | R |
| A2 | *A. baumanii* | Surg ward | R | R | R | I | R | R | R | R | I | I | I | I | S | R | R | S | R | R | S | S |
| A10 | *A. baumanii* | Med ward | R | R | R | I | R | R | R | R | R | R | R | R | R | R | I | I | R | R | S | R |
| A12 | *A. baumanii* | ICU | R | R | R | R | R | R | R | R | R | R | R | R | R | R | R | R | R | R | S | R |
| A3 | *A. baumanii* | Surg ward | R | R | R | R | R | R | R | R | R | R | R | R | R | R | R | R | R | R | R | R |
| A6 | *A. baumanii* | ICU | R | R | R | R | R | R | R | R | R | R | R | R | R | R | R | R | R | R | R | R |
| A8 | *A. baumanii* | ICU | R | R | R | R | R | R | R | R | R | R | R | R | R | R | R | R | R | R | R | R |
| A11 | *A. baumanii* | ICU | R | R | R | R | R | R | R | R | R | R | R | R | R | R | R | R | R | R | R | R |
| A9 | *A. baumanii* | POPD | R | R | R | R | R | R | R | R | R | R | R | R | R | R | R | I | R | R | S | R |
| A1 | *A. baumanii* | POPD | R | R | R | R | R | R | R | R | R | R | R | R | R | R | I | R | R | R | S | R |
| A4 | *A. baumanii* | ICU | R | R | R | I | I | R | R | R | I | I | I | I | S | S | R | I | R | I | S | S |
| P5 | *P. aeruginosa* | Burns ward | S | R | S | S | I | S | R | R | S | S | S | R | R | S | S | R | R | R | S | S |
| P6 | *P. aeruginosa* | Burns ward | R | R | R | S | I | S | R | R | S | S | S | R | S | S | S | R | R | R | S | R |
| P2 | *P. aeruginosa* | Surg ward | R | R | R | I | R | R | R | R | S | S | S | R | R | S | S | R | R | R | S | R |
| P7 | *P. aeruginosa* | Burns ward | R | R | R | R | I | R | R | R | I | S | S | R | S | R | S | R | I | R | S | R |
| P1 | *P. aeruginosa* | Surg ward | R | R | R | S | R | R | R | R | R | S | I | R | R | S | S | R | R | R | R | R |
| P3 | *P. aeruginosa* | POPD | R | R | R | S | I | S | R | R | S | S | S | R | S | S | S | R | R | R | R | R |
| P8 | *P. aeruginosa* | ICU | R | R | R | S | S | S | R | R | S | S | S | R | S | S | S | R | R | R | S | R |
| P4 | *P. aeruginosa* | Burns ward | R | R | R | S | R | S | R | R | S | S | S | R | S | S | S | R | R | R | S | R |

Abbreviations: AMP, ampicillin; AMC, amoxicillin–clavulanic acid; CTX, cefotaxime; CAZ, ceftazidime; CRO, ceftriaxone; FEP, cefepime; LEX, cephalexin; FOX, cefoxitin; TZP, piperacillin–tazobactam; IPM, imipenem; MEM, meropenem; NAL, nalidixic acid; CIP, ciprofloxacin; GEN, gentamicin; AMK, amikacin; TET, tetracycline; TGC, tigecycline; CHL, chloramphenicol, SXT, trimethoprim–sulfamethoxazole, and AZM, azithromycin. R, resistant; I, intermediate susceptibility; S, susceptible; ICU, Intensive Care Unit; Med ward, medical ward; POPD, paediatric outpatient department; Paed ward; Paediatric ward; Surg ward, Surgical ward;

### Table S5: Detailed phenotypic profiles of *S. aureus* isolates

| Isolate ID | Species | Ward type | PEN | AMP | FOX | CIP | MXF | LEV | GEN | AMK | TGC | CHL | NIT | SXT | ERY | LZD | TEC | TET | DO | RIF | CLI | VAN |
|---|---|---|---|---|---|---|---|---|---|---|---|---|---|---|---|---|---|---|---|---|---|---|
| S2 | *S. aureus* | Med ward | R | S | S | R | S | S | S | I | R | S | S | S | I | R | I | R | R | I | I | S |
| S3 | *S. aureus* | Emerg dept | R | S | S | S | S | S | S | S | R | R | R | I | I | R | I | I | I | S | I | S |
| S11 | *S. aureus* | Surg ward | R | R | R | R | R | R | R | I | R | S | S | R | 1 | S | I | R | R | R | R | S |
| S21 | *S. aureus* | Emerg dept | R | S | S | R | R | R | R | I | R | S | S | R | I | S | I | R | R | R | S | S |
| S22 | *S. aureus* | Med ward | R | S | S | S | S | S | S | S | R | S | S | S | I | S | I | I | I | I | I | S |
| S24 | *S. aureus* | Surg ward | R | R | R | S | R | S | S | S | R | R | R | R | R | R | R | R | R | R | R | S |
| S25 | *S. aureus* | Surg ward | R | S | S | R | S | R | R | I | S | S | S | R | I | S | I | R | R | I | S | S |
| S33 | *S. aureus* | Med ward | R | S | S | S | S | S | R | I | R | S | S | S | I | S | S | I | R | R | S | S |
| S38 | *S. aureus* | Med ward | R | S | S | S | S | S | S | S | R | S | S | S | I | S | S | S | S | I | S | S |
| S39 | *S. aureus* | Surg ward | R | R | R | R | S | S | R | S | S | S | S | S | S | S | S | R | R | I | S | S |
| S41 | *S. aureus* | Med ward | R | S | S | R | R | R | R | S | S | S | S | R | R | R | R | I | R | R | R | S |
| S42 | *S. aureus* | OOPD | R | R | R | R | S | R | R | I | R | S | S | I | R | S | I | S | R | I | S | S |
| S43 | *S. aureus* | Med ward | R | R | R | R | R | R | R | S | R | S | S | R | R | S | S | I | I | I | S | S |
| S49 | *S. aureus* | Emerg dept | R | S | S | R | R | R | S | I | R | S | S | S | I | S | I | I | I | S | I | S |
| S18 | *S. aureus* | Extn ward | R | R | R | R | R | R | R | R | R | S | R | R | R | R | R | R | R | R | S | S |
| S30 | *S. aureus* | Burns ward | R | S | S | R | S | S | S | S | S | S | S | S | S | S | I | I | S | S | S | S |
| S31 | *S. aureus* | Burns ward | R | R | R | R | R | I | R | R | S | S | I | R | R | S | R | R | R | I | S | S |
| S13 | *S. aureus* | Paed ward | R | S | S | S | S | R | S | S | S | S | I | R | R | R | I | R | S | R | S | S |
| S46 | *S. aureus* | Extn ward | R | R | R | S | S | S | I | S | S | S | I | S | S | S | I | I | S | S | S | S |
| S6 | *S. aureus* | ICU | R | S | S | R | R | I | R | S | R | S | R | R | R | I | R | R | S | I | S | S |
| S7 | *S. aureus* | OOPD | R | R | R | R | R | R | I | R | R | S | R | R | R | I | R | S | S | I | S | S |
| S12 | *S. aureus* | Paed ward | R | R | S | R | R | R | R | I | S | S | R | R | R | I | I | R | S | I | S | S |
| S15 | *S. aureus* | POPD | R | R | S | R | R | 1 | S | R | S | S | R | S | S | I | R | R | S | R | S | S |
| S19 | *S. aureus* | POPD | R | R | S | R | R | I | R | R | S | S | S | S | S | I | S | I | R | I | S | S |
| S28 | *S. aureus* | POPD | S | S | S | S | R | S | I | R | S | S | R | S | S | S | I | R | R | I | S | S |
| S29 | *S. aureus* | Paed ward | R | R | R | R | R | S | R | S | S | S | S | R | S | I | I | I | R | I | S | S |
| S32 | *S. aureus* | ICU | R | R | R | S | R | S | I | R | S | S | S | R | S | S | R | I | R | R | S | S |
| S34 | *S. aureus* | POPD | R | S | S | R | S | S | S | I | S | S | S | S | S | S | S | I | S | R | S | S |
| S40 | *S. aureus* | POPD | R | S | S | S | R | S | S | R | S | S | S | S | S | S | S | I | R | S | S | S |

| | | | | | | | | | | | | | | | | | | | | | | |
|---|---|---|---|---|---|---|---|---|---|---|---|---|---|---|---|---|---|---|---|---|---|---|
| S44 | *S. aureus* | ICU | R | R | R | R | R | R | S | R | S | S | R | R | R | R | R | R | R | R | S | S |
| S45 | *S. aureus* | OOPD | R | S | R | R | R | S | I | R | S | S | I | R | S | I | R | R | R | R | S | S |
| S48 | *S. aureus* | ICU | R | R | R | R | R | S | R | R | S | S | S | R | S | I | S | I | S | S | S | S |
| S1 | *S. aureus* | POPD | R | S | S | R | R | S | R | R | R | S | R | S | S | R | R | R | S | R | S | S |
| S4 | *S. aureus* | KMMC clinic | R | R | R | S | S | 1 | R | R | R | S | S | R | R | R | R | R | S | R | S | S |
| S5 | *S. aureus* | KMMC clinic | R | S | S | R | R | R | S | R | R | S | S | R | R | I | R | I | R | I | S | S |
| S8 | *S. aureus* | Emerg dept | R | S | S | R | R | R | R | I | S | S | R | R | R | I | R | I | S | R | S | S |
| S9 | *S. aureus* | POPD | S | S | S | S | S | R | R | R | R | S | S | R | R | R | R | I | S | R | S | S |
| S10 | *S. aureus* | POPD | R | S | S | R | S | I | R | R | R | S | S | R | S | R | I | R | R | I | S | S |
| S13 | *S. aureus* | POPD | R | R | R | R | R | R | S | I | R | S | I | R | R | R | R | R | R | I | S | S |
| S14 | *S. aureus* | OOPD | R | S | S | S | R | R | I | R | S | S | S | R | S | I | I | I | S | I | S | S |
| S16 | *S. aureus* | POPD | R | S | S | R | R | I | I | I | S | S | S | R | S | I | I | R | S | I | S | S |
| S17 | *S. aureus* | POPD | R | S | S | S | S | I | S | R | R | S | S | R | S | I | I | S | S | I | S | S |
| S20 | *S. aureus* | Surg ward | S | S | S | S | S | I | I | R | S | S | S | S | R | I | I | R | S | I | S | S |
| S26 | *S. aureus* | POPD | R | R | R | R | R | R | I | S | R | R | R | R | R | R | R | R | R | R | S | S |
| S37 | *S. aureus* | Paed ward | R | I | I | R | S | S | I | S | S | S | S | R | S | S | I | I | S | R | S | S |

Abbreviations: FOX, cefoxitin; PEN, penicillin G; CPT, ceftaroline; CIP, ciprofloxacin; MXF, moxifloxacin; AZM, azithromycin; ERY, erythromycin; GEN, gentamicin; AMK, amikacin; CHL, chloramphenicol; TET, tetracycline; DOX, doxycycline; TEC, teicoplanin; TGC, tigecycline; LZD, linezolid; CLI, clindamycin; RIF, rifampicin; SXT, sulphamethoxazole/trimethoprim; NIT, nitrofurantoin; R, resistant; OPD, Outpatient Department; ICU, Intensive Care Unit;Emerg dept; Emergency department, OOPD; Orthopedic out-patient department

**Table S6: Detailed phenotypic profiles of *E. faecium* isolates**

| Isolate ID | Species | Ward type | Antibiotic resistance profile | | | | | | | | | | | | | | | |
|---|---|---|---|---|---|---|---|---|---|---|---|---|---|---|---|---|---|---|
| | | | **AMP** | **IMP** | **CIP** | **LEV** | **GEN** | **STR** | **ERY** | **TGC** | **CHL** | **NIT** | **SXT** | **LZD** | **TEC** | **VAN** | **TET** | **QD** |
| EF10 | *E. faecium* | POPD | S | I | S | S | S | S | R | R | S | R | R | R | R | R | R | R |
| EF6 | *E. faecium* | ICU | R | R | R | R | R | R | I | S | I | S | R | R | S | S | I | R |
| EF5 | *E. faecium* | Med ward | R | R | R | R | R | S | R | I | S | S | R | S | S | S | R | R |
| EF7 | *E. faecium* | Med ward | R | R | R | R | R | S | I | S | S | R | R | S | S | S | 1 | S |
| EF8 | *E. faecium* | Paed ward | R | R | R | R | R | R | I | I | S | S | R | S | S | S | I | I |
| EF9 | *E. faecium* | Med ward | S | S | S | S | S | S | R | I | S | S | R | S | S | S | S | R |
| EF12 | *E. faecium* | POPD | S | S | S | S | S | R | I | I | R | S | R | R | S | S | R | R |
| EF13 | *E. faecium* | POPD | S | R | R | R | R | R | R | S | I | S | R | S | S | S | S | I |
| EF15 | *E. faecium* | POPD | R | R | R | R | R | R | R | I | I | S | R | S | S | S | R | R |
| EF19 | *E. faecium* | KMMC clinic | S | I | S | S | S | S | R | I | R | R | I | S | S | S | R | R |
| EF21 | *E. faecium* | Med ward | S | S | S | S | S | S | I | I | R | S | R | S | S | S | R | R |
| EF23 | *E. faecium* | ICU | R | R | R | S | S | S | R | S | R | R | I | S | S | S | R | R |
| EF24 | *E. faecium* | Surg ward | R | R | R | R | R | R | R | S | S | I | R | S | S | S | R | R |
| EF25 | *E. faecium* | KMMC clinic | R | R | R | R | R | R | R | I | S | S | R | S | S | S | I | R |
| EF26 | E. faecium | Paed ward | S | S | S | S | S | S | R | I | S | S | I | S | S | S | R | R |

PEN = penicillin G; AMP= ampicillin; AMC= amoxicillin-clavulanic acid; LEX= cephalexin; CTX= cefotaxime; CAZ=cefotaxime; FEP= cefepime; CRO= ceftriaxone; FOX = cefoxitin; TZP= piperacillin-tazobactam; IMP= imipenem; MEM= meropenem; CIP = ciprofloxacin; MXF = moxifloxacin; AZM = azithromycin; ERY = erythromycin; GEN = gentamicin; AMK = amikacin; CHL = chloramphenicol; TET = tetracycline; DOX = doxycycline; TEC = teicoplanin; TGC = tigecycline; LZD = linezolid; CLI = clindamycin; NAL= nalidixic acid; RIF = rifampicin; SXT = sulfamethoxazole/trimethoprim; NIT = nitrofurantoin. I, intermediate susceptibility; Q-D, quinupristin-dalfopristin; R, resistant; S, susceptible; VAN, vancomycin OOPD; Orthopedic out-patient department; Paed ward, Paediatric ward

**Table S7: Multiple antibiotic resistant phenotypes displayed by ESKAPE and *E. coli* isolates**

| Species | Resistance Phenotype | No. observed |
|---|---|---|
| ***E. faecium*** | AMP-IMP-CIP-GEN-ERY-SXT-TET-STR-LEV-QD | 2 |
| | ERY-TGC-NIT-SXT-LZD-TEC-TET-VAN-QD | 1 |
| | AMP-IMP-CIP-GEN-ERY-SXT-TET-LEV- QD | 1 |
| | AMP-IMP-CIP-GEN-ERY-SXT-STR-LEV- QD | 1 |
| | AMP-IMP-CIP-GEN-SXT-LZD-STR-LEV-QD | 1 |
| | IMP-CIP-GEN-ERY-SXT-STR-LEV | 1 |
| | AMP-IMP-CIP-GEN-STR-LEV | 1 |
| | CHL-SXT-LZD-TET-STR-QD | 1 |
| | ERY-CHL-NIT-TET-QD | 1 |
| | CHL-SXT-TET-QD | 1 |
| | ERY-SXT-QD | 1 |
| ***S. aureus*** | PEN-G-AMP-FOX-CIP-MXF-LEV-GEN-AMK-TGC-NIT-SXT-ERY-LZD-TEC-TET-DOX-RIF | 1 |
| | PEN-G-CIP-AMP-FOX-MXF-LEV-TGC-CHL-NIT-SXT-ERY-LZD-TEC-TET-DOX-RIF | 1 |
| | PENG-AMP-FOX-CIP-MXF-LEV-AMK-NIT-SXT-ERY-LZD-TEC-TET-DOX-RIF | 1 |
| | PEN-G-AMP-FOX-MXF-TGC-CHL-NIT-SXT-ERY-LZD-TEC-TET-DOX-RIF-CLI | 1 |
| | PEN-G-AMP-FOX-CIP-MXF-LEV-GEN-TGC-SXT-ERY-TET-DOX-RIF-CLI | 1 |
| | PENG-AMP-FOX-LEV-GEN-AMK-TGC-SXT-ERY-LZD-TEC-TET-RIF | 1 |
| | PENG-AMP-FOX-CIP-MXF-GEN-AMK-SXT-ERY-TEC-TET-DOX | 1 |
| | PEN G-AMP-FOX-CIP-MXF-LEV-TGC-SXT-ERY-LZD-TEC-TET-DOX | 1 |
| | PENG-CIP-MXF-LEV-GEN-SXT-ERY-LZD-TEC-DOX-RIF-CLI | 1 |
| | PENG-AMP-FOX-CIP-MXF-LEV-AMK-TGC-NIT-SXT-ERY-TEC | 1 |
| | PENG-AMP-CIP-MXF-LEV-GEN-NIT-SXT-ERY-TET | 1 |
| | PENG-AMP-CIP-MXF-LEV-AMK-NIT-TEC-TET-RIF | 1 |
| | PENG-AMP-FOX-CIP-MXF-AMK-SXT-TEC-TET-DOX-RIF | 1 |
| | PENG-CIP-MXF-GEN-AMK-TGC-NIT-LZD-TEC-TET-RIF | 1 |
| | PENG-AMP-FOX-CIP-GEN-AMK-TGC-SXT-LZD-TET-DOX | 1 |
| | PENG-CIP-MXF-LEV-GEN-TGC-SXT-TET-DOX-RIF | 1 |
| | PENG-AMP-FOX-CIP-MXF-LEV-GEN-TGC-SXT-ERY | 1 |

| | | |
|---|---|---|
| | PENG-CIP-MXF-GEN-TGC-NIT-SXT-ERY-TEC-TET | 1 |
| | PENG-CIP-MXF-LEV-AMK-TGC-SXT-ERY-TEC-DOX | 1 |
| | PENG-CIP-MXF-LEV-GEN-NIT-SXT-ERY-TEC-RIF | 1 |
| | PENG-AMP-FOX-CIP-LEV-GEN-TGC-ERY-DOX | 1 |
| | LEV-GEN-AMK-TGC-SXT-ERY-LZD-TEC-RIF | 1 |
| | PENG-AMP-FOX-MXF-AMK-SXT-TEC-DOX-RIF | 1 |
| | PENG-AMP-FOX-CIP-MXF-GEN-SXT-DOX | 1 |
| | PENG-AMP-FOX-CIP-MXF-GEN-AMK-SXT | 1 |
| | PENG-CIP-LEV-GEN-SXT-TET-DOX | 1 |
| | PENG-AMP-FOX-CIP-GEN-TET-DOX | 1 |
| | PENG-CIP-TGC-LZD-TET-DOX | 1 |
| | PENG-LEV-SXT-ERY-LZD-TET-RIF | 1 |
| | PENG-AMP-CIP-MXF-GEN-AMK-DOX | 1 |
| | PENG-CIP-MXF-LEV-TGC | 1 |
| | MXF-AMK-NIT-TET-DOX | 1 |
| | PENG-TGC-CHL-NIT-LZD | 1 |
| | PENG-GEN-TGC-DOX-RIF | 1 |
| | PENG-MXF-LEV-AMK-SXT | 1 |
| | PENG-CIP-MXF-SXT-TET | 1 |
| | PENG-AMK-TGC-SXT | 1 |
| | PENG-MXF-AMK-DOX | 1 |
| | PENG-CIP-SXT-RIF | 1 |
| | PENG-CIP-RIF | 1 |
| | AMK-ERY-TET | 1 |
| ***K. pneumoniae*** | AMC-CTX-TZP -CAZ-CRO-FEP-LEX-FOX -IMP-MEM-NAL-CIP-GEN-AMK-TGC-CHL-SXT-AZM-TET | 1 |
| | AMC-TZP-CTX-CAZ-CRO-FEP-LEX-FOX -MEM-NAL-CIP-GEN-AMK-TGC-CHL-SXT-AZM-TET | 2 |
| | AMC-TZP-CTX-CAZ-CRO-FEP-LEX-FOX -IMP-MEM-NAL-CIP-GEN-AMK-TGC-CHL-SXT-AZM | 2 |
| | AMC-TZP-CTX-CAZ-CRO-FEP-LEX-FOX-IMP-MEM-NAL-CIP-GEN-TGC-CHL-SXT-AZM-TET | 1 |
| | AMC-TZP-CTX-CAZ-CRO-FEP-LEX-FOX-NAL-CIP-GEN-AMK-TGC-CHL-SXT-AZM-TET | 1 |
| | AMC-TZP-CTX-CAZ-CRO-FEP-LEX-FOX-MEM-NAL-CIP-GEN-AMK-TGC-CHL-SXT-TET | 1 |
| | AMC-TZP-CTX-CAZ-CRO-FEP-LEX-FOX-IMP-MEM-NAL-CIP-GEN-AMK-CHL-SXT-AZM | 1 |
| | AMC-TZP -CTX-CAZ-CRO-FEP-LEX-FOX-NAL-CIP-GEN-AMK-TGC-CHL-SXT-AZM-TET | 1 |

| | | |
|---|---|---|
| | AMC-TZP -CTX-CAZ-CRO-FEP-LEX-FOX-MEM-NAL-CIP-GEN-TGC-CHL-SXT-AZM-TET | 1 |
| | AMC-TZP-CTX-CAZ-CRO-FEP-LEX-FOX-NAL-CIP-GEN-AMK-TGC-CHL-SXT-TET | 3 |
| | AMC-TZP-CTX-CAZ-CRO-FEP-LEX-FOX-IMP-NAL-CIP-GEN-AMK-TGC-CHL-SXT | 1 |
| | AMC-TZP-CTX-CAZ-CRO-FEP-LEX-FOX-IMP-MEM-NAL-CIP-GEN-TGC-SXT-TET | 1 |
| | AMC-TZP-CTX-CAZ-CRO-FEP-LEX-FOX-NAL-CIP-GEN-AMK-TGC-SXT-AZM-TET | 1 |
| | AMC-TZP-CTX-CAZ-CRO-FEP-LEX-FOX-NAL-CIP-GEN-AMK-TGC-CHL-SXT-TET | 1 |
| | AMC-TZP-CTX-CAZ-CRO-FEP-LEX-FOX-IMP-MEM-CIP-GEN-AMK-TGC-CHL-SXT | 1 |
| | AMC-TZP-CTX-CAZ-CRO-FEP-LEX-FOX-IMP-MEM-NAL-CIP-GEN-CHL-SXT-AZM | 1 |
| | AMC-TZP-CTX-CAZ-CRO-FEP-LEX-FOX-NAL-CIP-GEN-CHL-SXT-AZM-TET | 2 |
| | AMC-TZP-CTX-CAZ-CRO-FEP-LEX-FOX-NAL-CIP-GEN-TGC-SXT-AZM-TET | 1 |
| | AMC-TZP-CTX-CAZ-CRO-FEP-LEX-FOX-MEM-NAL-CIP-GEN-TGC-SXT-TET | 1 |
| | AMC-TZP-CTX-CAZ-CRO-FEP-LEX-FOX-NAL-CIP-GEN-CHL-SXT-TET | 1 |
| | AMC-CTX-CAZ-CRO-FEP-LEX-FOX-NAL-CIP-GEN-TGC-CHL-SXT-TET | 1 |
| | AMC-TZP -CTX-CAZ-CRO-FEP-LEX-FOX-NAL-CIP-GEN-TGC-CHL-SXT | 1 |
| | AMC-TZP-CTX-CAZ-CRO-FEP-LEX-FOX-NAL-CIP-GEN-AMK-TGC-SXT | 1 |
| | AMC-TZP -CTX-CAZ-CRO-FEP-LEX-FOX-MEM-NAL-CIP-GEN-SXT-TET | 1 |
| | AMC-TZP-CTX-CAZ-CRO-FEP-LEX-FOX-CIP-GEN-TGC-CHL-SXT-TET | 1 |
| | AMC-TZP -CTX-CAZ-CRO-FEP-LEX-FOX-NAL-CIP-GEN-CHL-SXT | 1 |
| | AMC-CTX-CAZ-CRO-FEP-LEX-FOX-NAL-CIP-GEN-TGC-SXT-TET | 1 |
| | AMC-CTX-CAZ-CRO-FEP-LEX-NAL-CIP-GEN-CHL-SXT-TET | 3 |
| | AMC-CTX-CAZ-CRO-FEP-LEX-FOX-NAL-CIP-GEN-TGC-SXT | 1 |
| | AMC-TZP-CTX-CAZ-CRO-FEP-LEX-FOX -NAL-CIP-GEN-SXT | 1 |
| | AMC-CAZ-CRO-FEP-LEX-FOX-GEN-TGC-CHL-AZM-TET | 1 |
| | AMC-TZP-CTX-CAZ-CRO-LEX-NAL-CIP-GEN-AMK-SXT | 1 |
| | AMC-TZP-CAZ-CRO-FEP-FOX -MEM-CIP-TGC-SXT-TET | 1 |
| | AMC-CRO-FEP-LEX-FOX-MEM-NAL-CIP-GEN-TGC | 1 |
| | AMC-TZP-CTX-CAZ-CRO-FEP-LEX -GEN | 1 |
| | AMC-TZP-FEP-FOX -CIP-TGC | 1 |
| ***A. baumanii*** | AMP-TZP -CTX-CAZ-CRO-FEP-LEX-FOX-IMP-MEM-NAL-CIP-GEN-AMK-CHL-SXT-AZM-TET | 3 |
| | AMP -AMC-TZP -CTX-CAZ-CRO-LEX-FOX-IMP-MEM-NAL-CIP-GEN-AMK-TGC-CHL-AZM-TET | 1 |
| | AMP-TZP-CTX-CAZ-CRO-FEP-LEX-FOX-IMP-MEM-NAL-CIP-GEN-AMK-CHL-SXT-AZM-TET | 1 |
| | AMC-TZP-CTX-CAZ-CRO-FEP-LEX-FOX-IMP-MEM-NAL-CIP-GEN-TGC-CHL-SXT-TET | 1 |
| | AMP-AMC-TZP-CTX-CAZ-CRO-FEP-LEX-FOX-IMP-MEM-NAL-CIP-GEN-AMK-SXT-TET | 1 |

| | | |
|---|---|---|
| | AMP-AMC-TZP-CTX-CRO-FEP-LEX-IMP-MEM-NAL-CIP-GEN-TGC-CHL-SXT-TET | 1 |
| | AMC-TZP-CTX-CAZ-CRO-FEP-LEX-FOX-IMP-MEM-NAL-CIP-TGC-CHL-SXT-TET | 1 |
| | AMP-AMC-FEP-FOX-GEN-AMK-SXT | 1 |
| | AMC-CTX-LEX-FOX-AMK-CHL | 1 |
| ***P. aeruginosa*** | AMP-AMC-TZP-CTX-CRO-FEP-LEX-FOX-NAL-CIP-TGC-CHL-SXT-AZM-TET | 1 |
| | AMP-AMC-CTX-LEX-FOX-NAL-TGC-CHL-SXT-TET | 3 |
| | AMP-AMC-CTX-CRO-FEP-LEX-FOX-NAL-CIP-TGC-CHL-SXT-TET | 1 |
| | AMP-AMC-CTXCAZ-FEP-LEX-FOX-NAL-GEN-TGC-SXT-TET | 1 |
| | AMP-AMC-CTX-LEX-FOX-NAL-TGC-CHL-SXT-AZM-TET | 1 |
| | AMC-LEX-FOX-NAL-CIP-TGC-CHL-SXT | 1 |
| ***Enterobacter* spp.** | AMP-AMC-TZP-CTX-CAZ-CRO-FEP- LEX-FOX-NAL-CIP-GEN-CHL-SXT- TET | 1 |
| | AMP-AMC-TZP-CTX-CAZ-CRO-LEX-FOX-SXT | 1 |
| | AMP-AMC-CTX-CAZ-CRO-FEP-LEX- FOX-SXT | 1 |
| | AMP-AMC-CTX-CRO-FEP-LEX-FOX-CIP-AZM | 1 |
| | AMP-AMC-CTX-CAZ-LEX-FOX-CIP-TGC | 1 |
| | AMP-AMC-CRO-LEX-FOX-CHL | 1 |
| | AMP-AMC-SXT-AZM-TET | 1 |
| | AMP-AMC-LEX-FOX-CIP | 1 |
| | AMP-CRO-CIP | 1 |
| ***E. coli*** | AMP-AMC-TZP-CTX-CAZ-CRO-FEP-LEX-FOX-IMP-MEM-NAL-CIP-GEN-AMK-TGC-CHL-SXT-AZM-TET | 1 |
| | AMP-AMC-CTX-CAZ-CRO-FEP-LEX-NAL-CIP-GEN-AMK-SXT-AZM-TET | 4 |
| | AMP-AMC-TZP-CTX-CAZ-CRO-FEP-LEX-FOX-NAL-CIP-GEN-SXT-AZM-TET | 2 |
| | AMP-AMC-TZP-CTX-CAZ-CRO-FEP-LEX-FOX-NAL-CIP-GEN-AMK-SXT-AZM-TET | 1 |
| | AMP-AMC-CTX-CAZ-CRO-FEP-LEX-FOX-NAL-CIP-GEN-CHL-SXT-AZM-TET | 1 |
| | AMP-AMC-TZP-CTX-CAZ-CRO-FEP-LEX-FOX-NAL-CIP-GEN-AMK-TGC -SXT | 1 |
| | AMP-AMC-CTX-CAZ-CRO-FEP-LEX-NAL-CIP-GEN-AMK-CHL-SXT-AZM-TET | 1 |
| | AMP-AMC-TZP-CTX-CAZ-CRO-FEP-LEX-FOX-CIP-GEN-AMK-TGC-SXT | 1 |
| | AMP-AMC-CTX-CAZ-CRO-FEP-LEX-NAL-CIP-GEN-CHL-SXT-AZM-TET | 1 |
| | AMP-AMC-TZP-CTX-CAZ-CRO-FEP-LEX-FOX-CIP-GEN-TGC-SXT | 1 |
| | AMP-AMC-CRO-LEX-FOX-IMP-MEM-NAL-CIP-CHL-SXT-AZM-TET | 1 |
| | AMP-AMC-CRO-LEX-FOX-IMP-MEM-CIP-GEN-AMK-CHL-SXT-TET | 1 |
| | AMP-AMC-CTX-CAZ-CRO-FEP-LEX-NAL-CIP-GEN-SXT-TET | 1 |

| | | |
|---|---|---|
| | AMP-AMC-TZP-CTX-CRO-NAL-CIP-AMK-CHL-SXT-TET | 1 |
| | AMP-AMC-CTX-CRO-LEX-NAL-CIP-SXT-AZM-TET | 1 |
| | AMP-AMC-TZP-CTX-CAZ-CRO-FEP-LEX -SXT | 1 |
| | AMP-AMC-NAL-CIP-CHL-SXT-AZM-TET | 1 |
| | AMP-AMC-FOX-CIP-SXT-AZM-TET | 1 |
| | AMP-AMC-GEN-SXT-AZM | 1 |

PEN = penicillin G; AMP= ampicillin; AMC= amoxicillin-clavulanic acid; LEX= cephalexin; CTX= cefotaxime; CAZ=cefotaxime; FEP= cefepime; CRO= ceftriaxone; FOX = cefoxitin; TZP= piperacillin-tazobactam; IMP= imipenem; MEM= meropenem; CIP = ciprofloxacin; MXF = moxifloxacin; AZM = azithromycin; ERY = erythromycin; GEN = gentamicin; AMK = amikacin; CHL = chloramphenicol; TET = tetracycline; DOX = doxycycline; TEC = teicoplanin; TGC = tigecycline; LZD = linezolid; CLI = clindamycin; NAL= nalidixic acid; RIF = rifampicin; SXT = sulfamethoxazole/trimethoprim; NIT = nitrofurantoin.

Table S8A: Results of Shapiro–Wilk and Levene's tests for normality and homogeneity of variances

| Comparison | Test | Statistic | P-value / Comment |
|---|---|---|---|
| WardGroup | Levene's Test | 1.001 | 0.427<br>Assumption met |
| Facility Level | Levene's Test | 3.303 | 0.071<br>Assumption met |

* Shapiro–Wilk tests indicated acceptable normality across groups for both Ward Group and Facility Level.

Table S8B: One-way ANOVA summary statistics for MARI values across ward groups and facility levels.

| Comparison | F-value | P-value | $\eta^2$ (Effect Size) | Post hoc |
|---|---|---|---|---|
| WardGroup | 2.895 | 0.011 | 0.103 | Tukey HSD: significant pairwise differences |
| Facility Level | 17.520 | < 0.001 | 0.101 | Not applicable<br>(2-group ANOVA) |

Table S8C: Tukey's HSD post hoc pairwise comparisons of MARI values among ward groups.

| Group 1 | Group 2 | Mean Difference | P-Value | Lower CI | Upper CI | Significant |
|---|---|---|---|---|---|---|
| Emergency/Trauma Units | General Inpatient Wards | -0.1394 | 0.6924 | -0.4024 | 0.1236 | False |
| Emergency/Trauma Units | Intensive Care Units | 0.0341 | 0.9994 | -0.1963 | 0.2645 | False |
| Emergency/Trauma Units | Outpatient/Clinic-Based Services | -0.1413 | 0.5767 | -0.3813 | 0.0986 | False |
| Emergency/Trauma Units | Paediatric Units | -0.1347 | 0.6813 | -0.386 | 0.1166 | False |
| Emergency/Trauma Units | Specialist Wards | -0.055 | 0.9975 | -0.3431 | 0.2331 | False |

| | | | | | | |
|---|---|---|---|---|---|---|
| Emergency/Trauma Units | Surgical Wards | 0.012 | 1.0 | -0.2482 | 0.2722 | False |
| General Inpatient Wards | Intensive Care Units | 0.1736 | 0.1278 | -0.0245 | 0.3717 | False |
| General Inpatient Wards | Outpatient/Clinic-Based Services | -0.0019 | 1.0 | -0.211 | 0.2072 | False |
| General Inpatient Wards | Paediatric Units | 0.0047 | 1.0 | -0.2174 | 0.2268 | False |
| General Inpatient Wards | Specialist Wards | 0.0844 | 0.9617 | -0.1786 | 0.3474 | False |
| General Inpatient Wards | Surgical Wards | 0.1515 | 0.4509 | -0.0807 | 0.3836 | False |
| Intensive Care Units | Outpatient/Clinic-Based Services | -0.1755 | 0.0313 | -0.3418 | -0.0092 | True |
| Intensive Care Units | Paediatric Units | -0.1689 | 0.0891 | -0.3512 | 0.0135 | False |
| Intensive Care Units | Specialist Wards | -0.0891 | 0.9091 | -0.3195 | 0.1413 | False |
| Intensive Care Units | Surgical Wards | -0.0221 | 0.9999 | -0.2165 | 0.1723 | False |
| Outpatient/Clinic-Based Services | Paediatric Units | 0.0066 | 1.0 | -0.1876 | 0.2008 | False |
| Outpatient/Clinic-Based Services | Specialist Wards | 0.0863 | 0.9344 | -0.1536 | 0.3263 | False |
| Outpatient/Clinic-Based Services | Surgical Wards | 0.1534 | 0.2864 | -0.0523 | 0.359 | False |
| Paediatric Units | Specialist Wards | 0.0797 | 0.9639 | -0.1716 | 0.331 | False |
| Paediatric Units | Surgical Wards | 0.1467 | 0.4163 | -0.0721 | 0.3655 | False |
| Specialist Wards | Surgical Wards | 0.067 | 0.9875 | -0.1932 | 0.3272 | False |

Table S9A: Descriptive statistics of MARI values by ward group, including mean, standard deviation, sample size, and 95% confidence intervals.

| WardGroup | Mean MARI | Sample Size | Standard Deviation | Standard Error | CI Lower (95%) | CI Upper (95%) |
|---|---|---|---|---|---|---|
| Emergency/Trauma Units | 0.6416 | 12 | 0.2438 | 0.0703 | 0.5036 | 0.7796 |
| General Inpatient Wards | 0.5022 | 18 | 0.2953 | 0.0696 | 0.3657 | 0.6386 |
| Intensive Care Units | 0.6758 | 43 | 0.2156 | 0.0328 | 0.6113 | 0.7402 |
| Outpatient/Clinic-Based Services | 0.5003 | 31 | 0.2143 | 0.0384 | 0.4248 | 0.5757 |
| Paediatric Units | 0.5069 | 23 | 0.2568 | 0.0535 | 0.4020 | 0.6119 |
| Specialist Wards | 0.5866 | 12 | 0.2208 | 0.0637 | 0.4616 | 0.7116 |
| Surgical Wards | 0.6536 | 19 | 0.2314 | 0.0530 | 0.5496 | 0.7577 |

Table S9B: Descriptive statistics of MARI values by facility level (regional vs tertiary), including mean, standard deviation, sample size, and 95% confidence intervals.

| Level | Mean MARI | Sample Size | Standard Deviation | Standard Error | CI Lower (95%) | CI Upper (95%) |
|---|---|---|---|---|---|---|
| REGIONAL | 0.5475 | 128 | 0.2420 | 0.0213 | 0.5055 | 0.5894 |
| TERTIARY | 0.7450 | 30 | 0.1855 | 0.0338 | 0.6785 | 0.8114 |
